# Cohort Profile: Investigating Antidepressant Response within Generation Scotland

**DOI:** 10.64898/2026.02.23.26346868

**Authors:** Megan Calnan, Amelia Edmondson – Stait, Hannah Milbourn, Esme Elsden, Anjali K. Henders, Emily L. Ball, Matthew H. Iveson, AMBER Research Team, AMBER Lived Experience Advisory Panel, Generation Scotland Team, Naomi R. Wray, Sonia Shah, Cathryn M. Lewis, Andrew M. McIntosh

**Author notes:** Correspondence: Andrew M McIntosh.

## Abstract

**Background:** The Antidepressant Medications: Biology, Exposure & Response (AMBER) research programme was established to investigate the biological mechanisms underlying antidepressant action and variability in treatment response. Generation Scotland holds detailed genomic, clinical, and health information with recontacting consent, making this cohort ideal for investigating these aims.

**Methods:** We deployed a questionnaire, developed with input from a Lived Experience panel, to the Generation Scotland cohort to gather data on their depressive symptoms, medication history, efficacy, and side effects to develop clinically meaningful phenotypes of antidepressant response. Invitations were sent to 15,117 Generation Scotland participants who were 18 years or older and consented to be recontacted. Between July and November 2025, 1,180 participants with a history of antidepressant treatment for depression completed the questionnaire.

**Results:** The sample was predominantly female (78.1%), self-identified as White (98.6%), and older (median age 57 years) than the wider Generation Scotland cohort (median 49 years) and Scottish population (median 41.3 years). Participants reported heterogeneous depressive symptom profiles spanning mood, anxiety, cognitive, sleep, behavioural, and physical domains. One-third of participants (31.1%) had taken three or more different antidepressants. Selective serotonin reuptake inhibitors (SSRIs) were the most common class (89.1%). Using self-reported treatment duration, discontinuation patterns, and efficacy, we developed a stringent classification system to capture treatment response extremes, where 23.8% were classified as responders and 1.5% as non-responders, with the majority unclassified.

**Conclusions:** Questionnaire data will be linked with electronic health records to validate antidepressant response classifications. Following validation, 25 responders and 25 non-responders will provide biological samples for DNA methylation profiling and generation of patient-derived cell lines. These models will be exposed to SSRIs to identify gene expression signatures and biological pathways distinguishing treatment response, integrating with genomic and clinical data across the AMBER project. These findings will provide a valuable resource for future antidepressant response research.

**Plain Language Summary:** Depression is a common mental health condition affecting millions of people worldwide. Antidepressant medications are the primary medication treatment, but response is highly variable with only about one-third of individuals achieving full symptom remission after their first medication trial. We don’t fully understand why some people respond well while others don’t. To help answer this question, the Antidepressant Medications: Biology, Exposure & Response (AMBER) research programme was established. This study utilised the Generation Scotland cohort, a large health study in Scotland. Between July and November 2025, we invited 15,117 Generation Scotland participants to complete a detailed questionnaire about their experiences with antidepressant medications. A total of 1,180 participants answered detailed questions about their depression symptoms, which medications they tried, how long they were on a medication, how well the medications worked, and what side effects they experienced. We found that people’s experiences with depression and antidepressants varied considerably. About one-third had tried three or more different antidepressants. Using strict criteria based on treatment duration, effectiveness ratings, and medication changes, we identified 281 people (24%) who responded very well to SSRIs (the most common type of antidepressant) and 18 people (1.5%) who did not respond despite trying multiple SSRIs. A key limitation is that all information was self-reported, so we will validate findings by linking questionnaire responses with medical records. In the future, we will collect blood samples from some participants to study the biological differences between responders and non-responders. This research will help us better understand why antidepressants work for some people but not others, which could lead to more personalised treatment approaches for depression.

## Why was the study set up?

Major depressive disorder (MDD) is a major public health burden and a leading cause of global disability (Global Burden of Disease Study, 2022; Patel et al., 2018). It is a highly clinically heterogeneous condition that likely groups individuals with different aetiologies (Cui et al., 2024; Marx et al., 2023). Pharmacological treatment for depression typically begins with selective serotonin reuptake inhibitors (SSRIs), which are recommended as the first-line antidepressant class in clinical prescribing guidelines (National Institute for Health and Care Excellence (NICE), 2022). Other commonly prescribed antidepressants include selective noradrenaline reuptake inhibitors (SNRIs), tricyclic antidepressants (TCAs), and atypical antidepressants (Shefler et al., 2025). However, treatment response is highly variable with only about one-third of individuals achieving full remission of depressive symptoms following their first antidepressant trial (Gaynes et al., 2009). Another one-third progress to treatment-resistant depression (TRD) (Thomas et al., 2013), defined as inadequate response to two or more different antidepressant treatments of appropriate dose and duration (European Medicines Agency, 2025; Fabbri et al., 2019; Iveson et al., 2025). This pattern highlights the substantial heterogeneity of MDD, its variable response to antidepressants, and the limitations of trial-and-error prescribing approaches.

Growing evidence suggests variation in antidepressant treatment response is influenced by genetic factors, with single nucleotide polymorphism (SNP)-based heritability estimates suggesting 13% of the variance in antidepressant response is attributable to common genetic variation (Pain et al., 2022). Identifying genetic factors underlying antidepressant response and non-response could enable earlier recognition of which individuals might benefit from first-line SSRIs and support the development of more personalised and effective treatments. However, progress in identifying genetic variants and biological pathways predicting treatment outcomes has been limited by challenges in defining antidepressant response and the lack of large-scale studies combining patient-reported experiences, prescribing information, and genetic data.

There is a critical need to collect antidepressant response data integrated with patient perspectives, longitudinal clinical data, and genetic information. Patient-reported outcomes are particularly valuable for capturing dimensions of treatment response that matter to patients themselves, yet few studies have incorporated this data into large-scale clinical and genetic research on antidepressant treatment response. The Antidepressant Medications: Biology, Exposure & Response (AMBER) project was developed to address these challenges.

The Antidepressant Medications: Biology, Exposure & Response (AMBER) study is a Wellcome Trust–funded project designed to investigate the biological mechanisms underlying antidepressant action and variability in antidepressant treatment outcomes. Through five interconnected workstreams, the AMBER research team leverages large-scale health datasets, including electronic health records (EHRs), and genetic and molecular information, to develop clinically meaningful and reproducible phenotypes of antidepressant response. By integrating patient-reported outcomes with EHR-derived metrics and molecular profiling, AMBER aims to establish more precise definitions of antidepressant response and non-response that can serve as a foundation for genomic discovery and guide the development of more effective, personalised treatments for depression (https://kcl.ac.uk/research/amber-antidepressant-medications-biology-exposure-response).

This paper presents work from one stream of AMBER, which focuses on collecting patient-reported antidepressant exposure and response within the Generation Scotland cohort. A bespoke questionnaire was developed to collect detailed self-reported data on depressive symptoms, antidepressant use, perceived efficacy, and side effects. Responses to the questionnaire were used to develop a classification of antidepressant response, which will be validated through linkage with participants’ EHRs. From those whose self-reported and EHR-derived classifications align, 25 SSRI responders and 25 SSRI non-responders will be invited to provide biological samples (blood and saliva). Both blood and saliva samples will undergo DNA extraction and methylation profiling to characterise genome wide epigenetic patterns associated with SSRI treatment response. Blood samples will also be used to generate patient-derived cell lines. These cell lines will be treated with SSRIs *in vitro* and gene expression changes in response to SSRI exposure will be compared between responders and non-responders. Identifying genes and pathways that differ between these groups will help uncover molecular mechanisms relevant to SSRI treatment response. This integrated approach will allow for a more precise and biologically grounded understanding of antidepressant response and non-response. This dataset combining patient-reported outcomes, validated phenotypes, and molecular profiles will provide a valuable resource for future studies investigating antidepressant treatment response.

## Who is in the sample?

Generation Scotland (GS) was established in 2006 as a consented, longitudinal, family-based cohort through a multi-institutional collaboration between Scottish universities and NHS partners (https://genscot.ed.ac.uk). It is a national, population-based resource designed to advance understanding of the genetic, environmental, and social determinants of health and disease. More than 24,000 participants were originally recruited via general practices across Scotland and provided extensive data through questionnaires, clinical assessments, and biological samples. The participants consented to linkage with their NHS EHRs and to being recontacted for future studies (Smith et al., 2013). To expand the cohort, Next Generation Scotland was launched in 2022 to build upon the original GS resource, bringing the total cohort to over 40,000 participants (Milbourn et al., 2024). GS includes comprehensive genetic, health, lifestyle, and sociodemographic data, including genotyping information, mental and physical health records, prescription data, and longitudinal follow-up through NHS linkage (Mitchell et al., 2025). This scope of information allows detailed investigation of how genetic and environmental factors contribute to disease risk, treatment response, and health outcomes over time. GS offers a uniquely powerful environment to study antidepressant response, as it contains genomic data, prescription histories, longitudinal health records, and an established mechanism for recontacting participants. This infrastructure enabled us to invite a large sample and identify individuals with previous antidepressant exposure, collect in-depth patient-reported data, and later validate treatment history and response classifications through EHR linkage.

The GS team managed the recruitment process and began sending invitations to take the questionnaire in July 2025. Invitations were sent to 15,117 GS and Next GS participants (13,952 by email and 1,165 by SMS) who were 18 or older, registered on the portal, and provided consent to be recontacted for future studies. The questionnaire was hosted on the online NextGen participant portal. Participants were notified about the study through email and SMS notifications, each containing information about the study and a secure link to the questionnaire. After logging into the portal, participants could review the participant information sheet and provide consent before proceeding.

A total of 1,771 participants consented to take part but were screened out because they had not taken an antidepressant for mood-related reasons and therefore could not proceed to the full questionnaire. Only those who indicated use of an antidepressant for mood-related indications were able to complete the questionnaire. From 1 July 2025 to 11 November 2025, a total of 1,257 participants started the questionnaire. Of these, 1,180 participants (93.9%) completed the questionnaire and self-reported taking at least one antidepressant for depression, forming the analytic sample for the present study. The remaining participants started but did not complete the survey. Figure 1 illustrates the study workflow.

**Figure 1.**
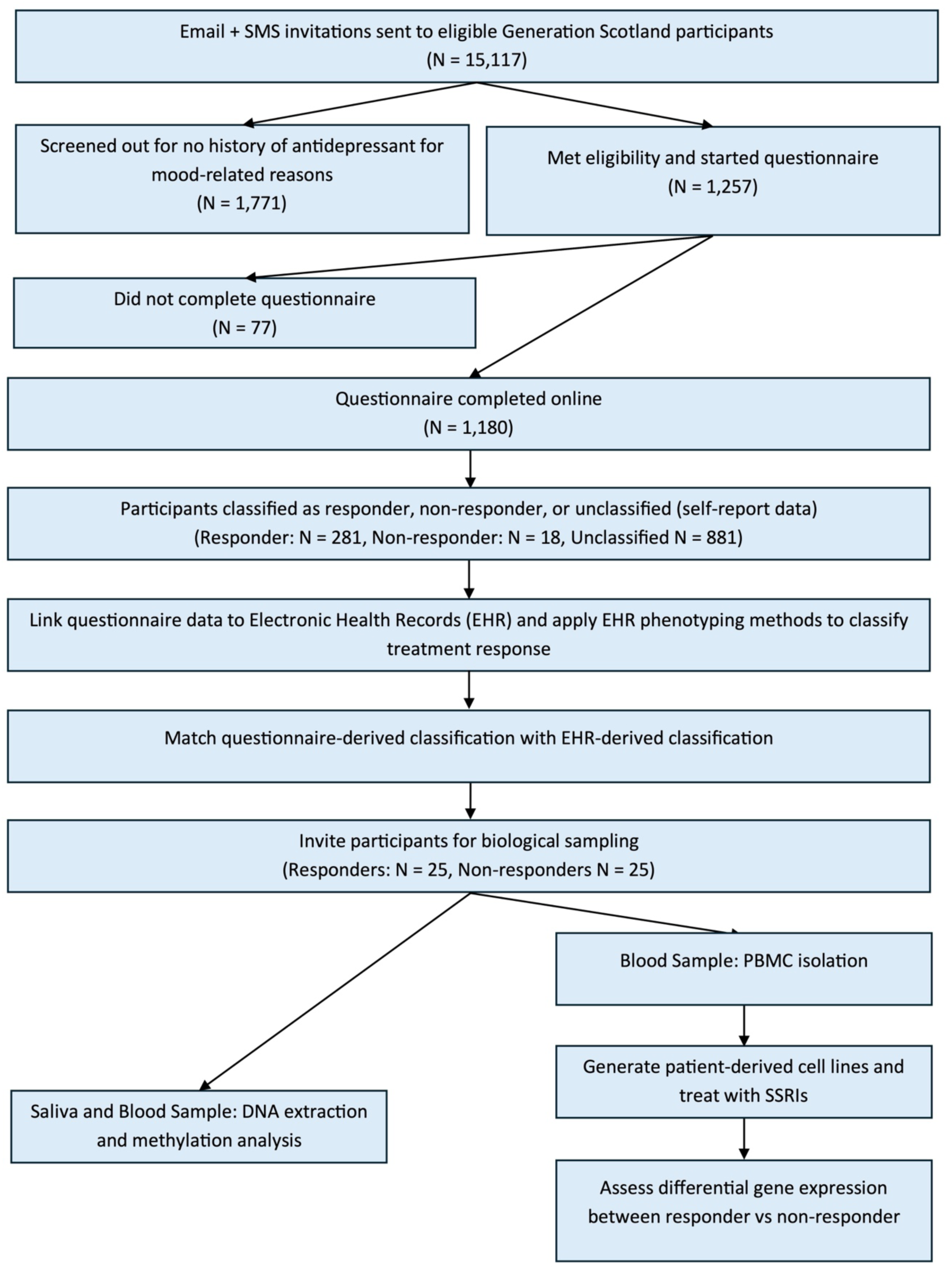
Overview of the study workflow. Flowchart showing participant recruitment, questionnaire completion, response classification, and future data collection plans. Email and SMS invitations were sent to 15,117 eligible Generation Scotland participants aged ≥18 who consented to recontact. Of 1,257 participants who started the questionnaire, 1,180 completed it online. Using self-reported treatment duration, discontinuation patterns, and perceived efficacy, participants were classified as SSRI responders (n=281), non-responders (n=18), or unclassified (n=881). Future workflow includes linking questionnaire data to Electronic Health Records (EHRs) for validation, matching questionnaire-derived and EHR-derived classifications, and inviting 25 responders and 25 non-responders for biological sampling. Blood samples will be used for peripheral blood mononuclear cell (PBMC) isolation to generate patient-derived cell lines for SSRI treatment and gene expression analysis. Saliva and blood samples will undergo DNA extraction for methylation analysis.

### Demographic Data

Demographic characteristics for this analytic sample were obtained from GS and Next GS baseline data and are summarised in comparison with the wider GS cohort and national estimates in Scotland from the 2011 census and 2022 population estimates in Table 1. Participants in this sample had a median age of 57 years. A majority fell within the 51-70 year age group (58.3%), followed by 31-50 years (25.4%), and 18 to 30 years was the least represented age range (7.5%). The analytic sample was predominantly female (78.1%), and most participants identified as White (98.6%).

**Table 1.** Participant Demographics (N = 1,180). Demographic characteristics of the analytic sample compared to the broader Generation Scotland cohort and Scottish population. Data include age, sex, ethnicity, geographic region, Scottish Urban Rural Classification, Scottish Index of Multiple Deprivation (SIMD), and depression/anxiety symptom severity measured by PHQ-9 (Patient Health Questionnaire-9) and GAD-7 (Generalized Anxiety Disorder-7) scales. SD = standard deviation.

| Covariate | Analytic Sample N | % | Generation Scotland % | Scottish Population % |
| --- | --- | --- | --- | --- |
| **Age (years)** |  |  |  |  |
| Mean (SD) | 54.3 (13.6) |  |  |  |
| Median | 57 |  | 49 | 41.3<sup>a</sup> |
| **Age group (years)** |  |  |  |  |
| 18 – 30 (%) | 88 | 7.5 |  |  |
| 31 – 50 (%) | 300 | 25.4 |  |  |
| 51 – 70 (%) | 688 | 58.3 |  |  |
| 71+ (%) | 104 | 8.8 |  |  |
| **Sex** |  |  |  |  |
| Female | 922 | 78.1 | 58.8 | 51.4<sup>a</sup> |
| Male | 249 | 21.1 | 41.2 | 48.6<sup>a</sup> |
| Missing | 9 | 0.8 |  |  |
| **Ethnicity** |  |  |  |  |
| White | 1,164 | 98.6 | 94.8 | 92.9<sup>a</sup> |
| Other | 14 | 1.2 | 1.1 | 7.1<sup>a</sup> |
| Missing | 2 | 0.2 | 4.1 |  |
| **Region** |  |  |  |  |
| Glasgow and Strathclyde | 320 | 27.1 | 33.5 | - |
| Tayside, Central and Fife | 248 | 21.0 | 36 | - |
| Edinburgh and Lothians | 246 | 20.8 | 10.8 | - |
| Aberdeen and North East | 162 | 13.7 | 11.9 | - |
| Highland and Islands | 136 | 11.5 | 5.5 | - |
| Scotland South | 63 | 5.3 | 2.4 | - |
| Missing | 5 | 0.4 | - | - |
| **Scottish Urban Rural Classification** |  |  |  |  |
| Large Urban Areas | 349 | 29.6 | 30.6 | 37.8<sup>b</sup> |
| Other Urban Areas | 370 | 31.4 | 27.7 | 33.9<sup>b</sup> |
| Accessible Small Towns | 123 | 10.4 | 8.4 | 8.6<sup>b</sup> |
| Remote Small Towns | 561 | 4.7 | 4.2 | 2.6<sup>b</sup> |
| Accessible Rural Areas | 185 | 15.7 | 11.1 | 11.6<sup>b</sup> |
| Remote Rural Areas | 92 | 7.8 | 5.8 | 5.5<sup>b</sup> |
| Missing | 5 | 0.4 | 12.2 | - |
| **Scottish Index of Multiple Deprivation** |  |  |  |  |
| Most Deprived Quintile | 133 | 11.3 | 11.3 | 19.2<sup>c</sup> |
| 2nd Most Deprived Quintile | 199 | 16.9 | 12.4 | 19.3<sup>c</sup> |
| Median Quintile | 267 | 22.6 | 14.3 | 19.8<sup>c</sup> |
| 2nd Least Deprived Quintile | 267 | 22.6 | 22.5 | 21.1<sup>c</sup> |
| Least Deprived Quintile | 309 | 26.2 | 27.3 | 20.6<sup>c</sup> |
| Missing | 5 | 0.4 | 12.2 | - |
| **PHQ-9** | 563 | 47.7 | - | - |
| PHQ-9 score, mean (SD) | 9.31 (7.23) | - | - | - |
| Minimal (0-4) | 181 | 32.1 | - | - |
| Mild (5-9) | 151 | 26.8 | - | - |
| Moderate (10-14) | 99 | 17.6 | - | - |
| Moderately Severe (15-19) | 63 | 11.2 | - | - |
| Severe (20-27) | 69 | 12.3 | - | - |
| Missing | 617 | 52.3 |  |  |
| **GAD-7** | 590 | 50.0 | - | - |
| GAD-7 score, mean (SD) | 7.36 (6.10) | - | - | - |
| Minimal (0-4) | 237 | 40.2 | - | - |
| Mild (5-9) | 176 | 29.8 | - | - |
| Moderate (10-14) | 71 | 12.0 | - | - |
| Severe (15-21) | 106 | 18.0 | - | - |
| Missing | 590 | 50.0 |  |  |

Generation Scotland Estimates based on their updated cohort profile output (Milbourn et al., 2024).
<sup>a</sup> Based on the 2011 Scottish Consensus (National Records of Scotland, 2023)
<sup>b</sup> Based on the estimates from Scottish Government Urban Rural Classification from NRS (National Records of Scotland., 2022).
<sup>c</sup> Based on the estimates from Population Estimates by SIMD from National Records of Scotland (NRS) (National Records of Scotland, 2022).

We compared demographics with both the wider GS cohort and national population estimates. The questionnaire respondents were older relative to the GS cohort (median age 49 years) (Milbourn et al., 2024) and national estimates (median age 41.3 years) (National Records of Scotland, 2023). Our sample also had higher proportion of female participants compared with the wider GS cohort (58.8%) and the national Scottish population (51.4%). The proportion of participants identifying as White was slightly higher compared to the wider GS cohort (94.8%) and Scottish census figures (92.9%).

Participants in our sample were distributed across all regions of Scotland, with the largest proportions located in Glasgow and Strathclyde (27.1%), Tayside, Central and Fife (21.0%), and Edinburgh and the Lothians (20.8%), and the smallest proportions in the southern regions of Scotland (5.3%). The Scottish Government Urban Rural Classification 2022 (Scottish Government, 2022) indicated the highest percentage of participants (31.5%) resided in other urban areas with populations of 10,000 to 124,999 and 7.8% resided in remote rural regions. Our sample had more representation in other urban areas (31.4%) compared to the entire GS cohort (27.7%), but less than the average in Scotland (33.9%). Lastly, our sample had a higher percentage of those in remote rural areas (7.8%) compared to GS (5.8%) and the national average (5.8%) (National Records of Scotland, 2022).

Socioeconomic diversity was captured using the Scottish Index of Multiple Deprivation (Scottish Government, 2020), with participants represented across all quintiles, 11.3% in the most deprived quintile and 26.2% in the least deprived quintile. This sample reflected equal representation to GS in the most deprived quintile (11.3%) but remained below the national average (19.2%). The proportion in the least deprived quintile within this sample (26.2%) was broadly comparable to GS (27.1%) and higher than national levels (20.6%) (National Records of Scotland, 2022).

## What has been measured?

### Depressive Symptoms

Patient Health Questionnaire (PHQ-9) and Generalised Anxiety Disorder (GAD-7) data were collected at the time of each participant’s original enrolment, prior to the questionnaire, and were used here to characterise depression and anxiety symptom severity within the cohort. Among antidepressant users with available data, 47.7% (n=563) of participants completed the PHQ-9 and 50.0% (n= 590) completed the GAD-7 (Table 1).

The mean (SD) PHQ-9 score was 9.31 (7.23; range 0-27), corresponding on average to mild-to-moderate depressive symptom severity (Kroenke et al., 2001). Using standard thresholds, 67.9% met criteria for any depression (PHQ-9 ≥ 5) and 41.0% for moderate-to-severe depression (PHQ-9 ≥ 10). The mean (SD) GAD-7 score was 7.36 (6.10; range 0-21), indicating mild anxiety symptom severity on average (Spitzer et al., 2006). 59.8% scored above the threshold for any anxiety (GAD-7 ≥ 5) and 30.0% met criteria for moderate-to-severe anxiety (GAD-7 ≥ 10).

PHQ-9 and GAD-7 scores were used descriptively to characterise depressive and anxiety symptoms among antidepressant users with available data from GS. These measures were collected at an earlier time point relative to the questionnaire utilised in this study, and scores should be interpreted in the context of ongoing, previous, or future antidepressant treatment, or periods of remission between depressive episodes. Lower scores may therefore reflect treatment response rather than the absence of symptoms, and participants may have been currently taking antidepressants, may have discontinued treatment, or may have initiated treatment after completing these earlier measures.

### Antidepressant Questionnaire

The questionnaire deployed to GS participants was adapted from the instrument developed in the Cell-omics Resource of the Australian Genetics of Depression Study (AGDS:Cell-o) (Mitchell et al., 2025). The questionnaire was designed for those with a history of antidepressant use for depression to collect detailed self-reported information on individual depressive symptoms, medication exposure, perceived efficacy, and side effects. The questionnaire was specifically created to fill a gap in capturing nuanced, multidimensional self-report data on people’s experience of depression and their antidepressant treatment journey and response. This questionnaire formed the foundation for developing the antidepressant response classification, which distinguishes between responders and non-responders based on detailed self-reported treatment histories.

To enhance its relevance and accessibility, the questionnaire was reviewed in collaboration with the AMBER study Lived Experience Advisory Panel (LEAP). The LEAP consists of individuals with personal experience of depression and antidepressant use, who provided critical feedback on the questionnaire’s wording, content, user-friendliness, and participant burden. Their input informed refinements to ensure that questions were clear, sensitive, and reflective of real-world experiences of antidepressant treatment. This co-production approach strengthened the validity and acceptability of the questionnaire among the target population.

The questionnaire included modules covering individual depressive symptoms and their impact on daily life, use of psychological and physical treatments for depression, antidepressant medication history, treatment duration, reasons for discontinuation, the perceived efficacy, and side effects of each antidepressant. A detailed range of depressive symptoms were captured across six domains: mood, anxiety, cognitive function, sleep, behaviour, and physical symptoms. For each selected symptom, participants rated the impact on their daily life from 1 (no impact) to 5 (severe impact).

Participants were presented with a list of the 15 most prescribed antidepressants in Scotland, along with their associated brand names, and asked whether they had been taken for mood-related symptoms. This served as a self-reported indicator of depression or related symptoms warranting antidepressant use. Participants were asked to indicate which medications they were currently taking or had taken in the past. For each reported antidepressant, participants provided details on duration of use, reason for discontinuation (if applicable), and perceived efficacy in reducing symptoms of depression. Participants also rated the medication’s impact in reduction of symptoms across the six symptom domains, providing a multidimensional profile of perceived treatment response. To capture time-to-response, participants were asked, *“How long did it take for you to notice any improvement in your symptoms after starting the medication(s)?”*

To align with other cohorts, we included the item: “Overall, do you feel [medication name, e.g., Citalopram or Celexa] is effective in reducing your symptoms of depression?” with response options: “Yes, a lot,” “Yes, a little,” or “No.” Finally, participants were asked whether they had experienced any of a predefined list of common side effects while taking each antidepressant, including nausea, dizziness, sleep disturbances, weight gain or loss, changes in appetite, fatigue, increased anxiety or agitation, sexual function or interest, headaches, feeling and being sick, indigestion or stomach aches, diarrhoea or constipation, dry mouth, memory problems, attention/concentration difficulties, shaking, muscle pain, fast heartbeat, itching/rash, and blurred vision. Participants were able to skip any questions they did not wish to answer. Figure 2 illustrates the modules included in the questionnaire.

**Figure 2.**
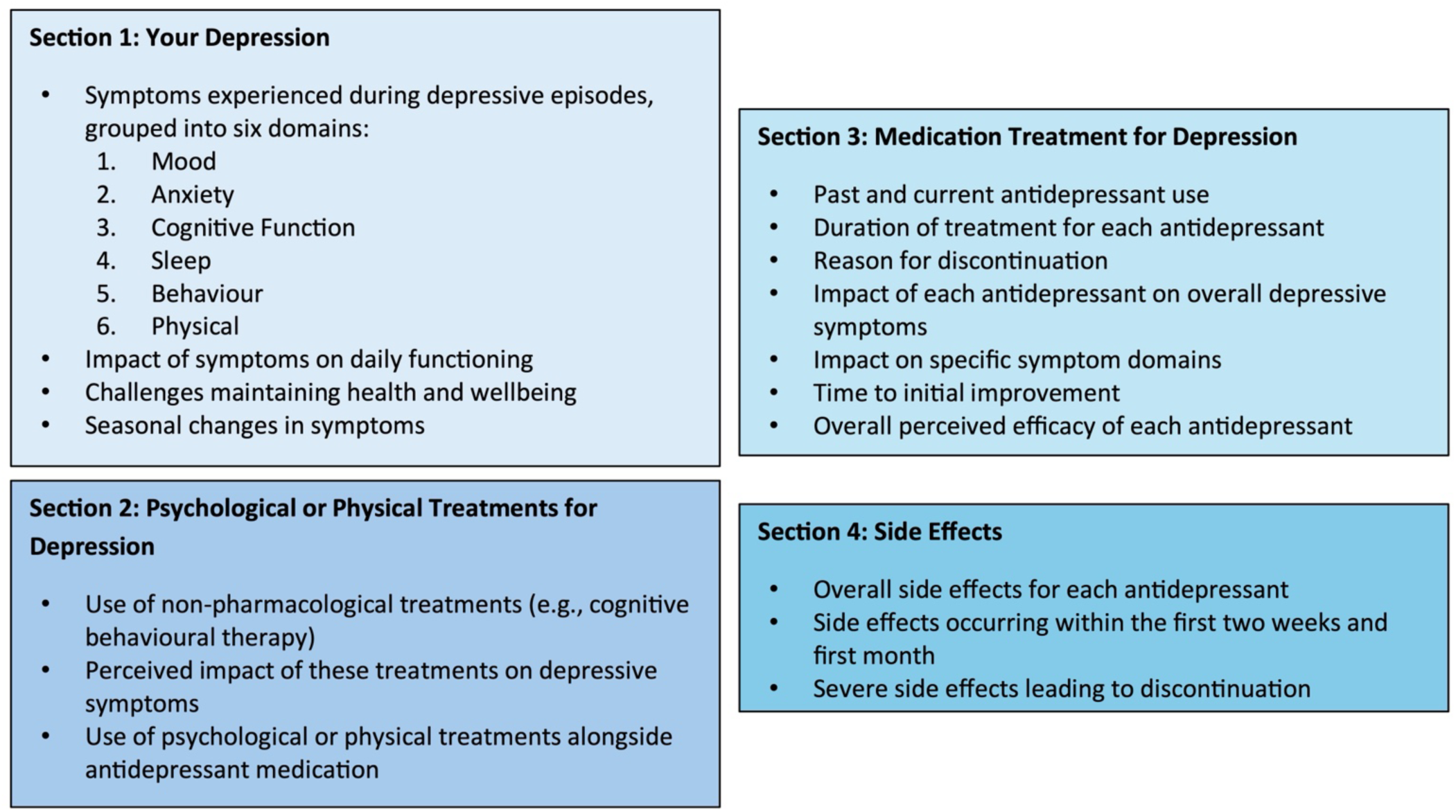
Overview of modules included in the questionnaire. Schematic showing the four main sections of the questionnaire. Section 1 (Your Depression) assessed symptoms experienced during depressive episodes across six domains: mood, anxiety, cognitive function, sleep, behaviour, and physical symptoms. Participants also reported symptom impact on daily functioning, challenges maintaining health and wellbeing, and seasonal changes in symptoms. Section 2 (Psychological or Physical Treatments for Depression) captured use of non-pharmacological treatments (e.g., cognitive behavioural therapy), their perceived impact on depressive symptoms, and whether these treatments were used alongside antidepressant medication. Section 3 (Medication Treatment for Depression) collected detailed information on past and current antidepressant use, including treatment duration, reason for discontinuation, impact on overall depressive symptoms and specific symptom domains, time to initial improvement, and overall perceived efficacy of each antidepressant. Section 4 (Side Effects) documented overall side effects for each antidepressant, side effects occurring within the first two weeks and first month of treatment, and severe side effects leading to discontinuation.

## What has it found?

All analyses were conducted using R (Version 4.4.1, R Core Team, 2024). Descriptive statistics were used to summarise participant demographics, antidepressant use, treatment response, and depressive symptom measures from both the questionnaire and linked PHQ-9 and GAD-7 data at GS enrolment. Categorical variables were reported as frequencies and percentages, and continuous variables as means with standard deviations (SD). The antidepressant response classification was operationalised using a rule-based framework derived from questionnaire responses on treatment duration, reasons for discontinuation, and perceived efficacy. Logical conditions were applied to assign participants into responder, non-responder, or unclassified groups. Missing data were excluded from relevant analyses on a per-variable basis.

### Antidepressant Questionnaire Depression Symptom Domains

The questionnaire assessed a range of depressive symptoms across six domains: mood, anxiety, cognitive functioning, sleep, behaviour, and physical symptoms. For each domain, participants were asked: *“When you are in a depressive episode, do you experience the following symptoms?”* If a symptom was endorsed, participants rated its impact on their daily life using a five-point scale from 1 (no impact) to 5 (severe impact). Figure S1 illustrates the questionnaire symptom domains assessed and the specific symptoms included within each domain.

The most endorsed symptoms across all domains were lack of energy (95.1%), waking unrefreshed/always fatigued (93.0%), reduced social activity (90.8%), loss of interest in hobbies (88.6%), and increased sensitivity to criticism or rejection (88.3%). These symptoms spanned multiple domains, reflecting the multidimensional nature of the cohort’s depressive symptomatology. Table 2 presents the 20 most endorsed symptoms alongside their mean impact ratings. Complete prevalence and impact data for each symptom by domain are available in Tables S1-6.

**Table 2.** The 20 most endorsed depressive symptoms with mean impact ratings on daily functioning. The 20 most frequently endorsed depression symptoms across six domains (Physical, Sleep, Behaviour, Mood, Anxiety, and Cognitive Function). Prevalence shows the number and percentage of participants endorsing each symptom. Impact ratings were measured on a 5-point scale (1 = no impact to 5 = severe impact on daily functioning). Symptoms are ordered by endorsement frequency, with lack of energy being the most prevalent (95.1% of participants endorsing). SD = standard deviation.

| Domain | Symptom | Prevalence<br>N (%) | Impact<br>Mean (SD) |
| --- | --- | --- | --- |
| Physical | Lack of energy | 1121 (95.1) | 3.66 (0.93) |
| Sleep | Waking unrefreshed/always fatigued | 1096 (93.0) | 3.78 (0.90) |
| Behaviour | Reduced social activity | 1068 (90.8) | 3.34 (0.95) |
| Depression | Loss of interest in hobbies | 1028 (88.6) | 3.44 (0.92) |
| Behaviour | Increased sensitivity to criticism/rejection | 1038 (88.3) | 3.50 (0.98) |
| Cognitive<br>Function | Everything feels like a burden | 1028 (87.4) | 3.60 (0.91) |
| Depression | Depressed mood: sadness, hopelessness,<br>frequent crying | 1018 (87.0) | 3.50 (0.86) |
| Cognitive<br>Function | Racing thoughts/can't shut off | 1010 (86.0) | 3.69 (0.97) |
| Anxiety | Anxious/worrying about minor things | 1004 (85.2) | 3.41 (0.93) |
| Cognitive<br>Function | Can't concentrate on book/movie | 983 (83.7) | 3.12 (0.96) |
| Depression | Worthlessness and self-doubt | 974 (83.6) | 3.54 (0.91) |
| Sleep | Waking throughout the night | 966 (82.1) | 3.48 (0.91) |
| Cognitive Function | Reduced ability to think/process | 942 (80.4) | 3.44 (0.89) |
| Cognitive Function | Can't concentrate at work/study | 936 (79.7) | 3.48 (0.93) |
| Cognitive Function | Decreased creativity | 915 (78.5) | 2.93 (1.00) |
| Behaviour | Reduced daytime activity | 911 (77.6) | 3.29 (0.87) |
| Sleep | Difficulty getting to sleep | 908 (77.1) | 3.66 (0.90) |
| Cognitive Function | Reduced ability to make decisions | 907 (77.4) | 3.46 (0.90) |
| Cognitive Function | Can't complete everyday tasks | 898 (76.5) | 3.52 (0.92) |
| Behaviour | Loss of interest in sex | 895 (77.4) | 2.99 (1.24) |

### Antidepressant Usage

Among the analytic sample (n = 1,180), participants reported a lifetime total of 2,674 antidepressants, with an average of 2.27 antidepressants per person (SD = 1.66). Just under half (n = 537, 45.5%) reported taking only one antidepressant, while 31.1% (n = 367) had taken three or more different antidepressants. The most common medications were the SSRIs: fluoxetine (n = 556, 47.1% of participants), sertraline (n= 506, 42.9%), and citalopram (n= 492, 41.7%). These were followed by the tricyclic antidepressant amitriptyline (n= 272, 23.1%) and the SNRI venlafaxine (n= 197, 16.7%). Table 3a displays all antidepressants including the number and proportion of participants who had taken each one. Note that many participants took more than one antidepressant.

**Table 3a.** Reported antidepressant use. Note: Many participants took more than one antidepressant. Frequency of antidepressant use by medication and drug class. Many participants took more than one antidepressant over their lifetime. Medications are ordered by frequency of use, with fluoxetine being the most commonly reported (47.1% of participants). SSRI = selective serotonin reuptake inhibitor; SNRI = serotonin-norepinephrine reuptake inhibitor.

| Antidepressant | Class | N (%) |
| --- | --- | --- |
| Fluoxetine | SSRI | 556 (47.1) |
| Sertraline | SSRI | 506 (42.9) |
| Citalopram | SSRI | 492 (41.7) |
| Amitriptyline | Tricyclic | 272 (23.1) |
| Venlafaxine | SNRI | 197 (16.7) |
| Mirtazapine | Atypical | 191 (16.2) |
| Paroxetine | SSRI | 159 (13.5) |
| Duloxetine | SNRI | 97 (8.2) |
| Escitalopram | SSRI | 56 (4.7) |
| Trazodone | Atypical | 37 (3.1) |
| Nortriptyline | Tricyclic | 30 (2.5) |
| Clomipramine | Tricyclic | 28 (2.4) |
| Lofepramine | Tricyclic | 23 (1.9) |
| Imipramine | Tricyclic | 20 (1.7) |
| Dosulepin | Tricyclic | 10 (0.8) |

SSRIs were the most prevalent drug class, with five agents accounting for a combined 1,051 users (89.1% of the sample). Tricyclic antidepressants were reported by 332 participants (28.1%), while SNRIs were reported by 249 participants (21.1%), and atypical antidepressants by 209 participants (17.7%). This distribution aligns with prescribing guidance recommended by the National Institute for Health and Care Excellence (NICE), which recommends SSRIs as the preferred first-line pharmacological treatment for depression in the United Kingdom (National Institute for Health and Care Excellence (NICE), 2022). Table 3b shows antidepressant usage by drug class, including the number and proportion of participants who reported each class.

**Table 3b.**
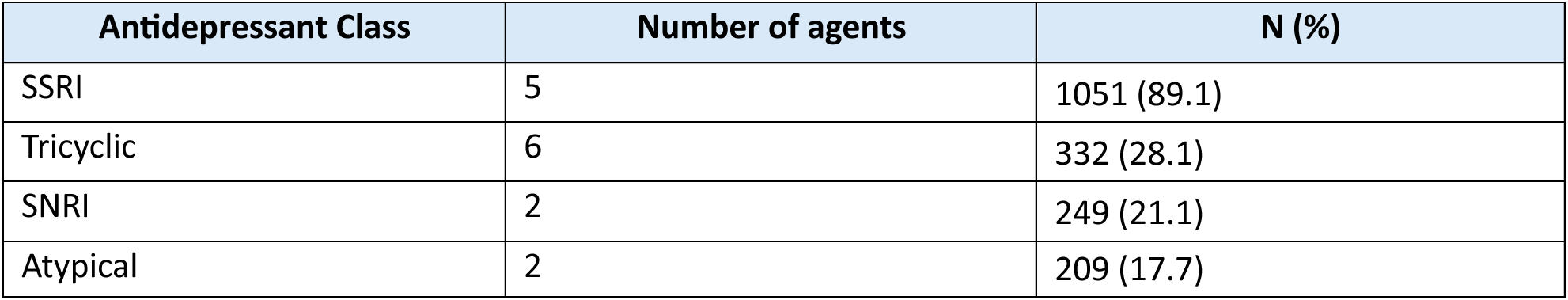
Summary of antidepressant use by drug class. Number of distinct antidepressant agents and total participants who used each drug class. SSRIs were the most common antidepressant class (89.1% of participants). Percentages sum to more than 100% as many participants used medications from multiple classes. SSRI = selective serotonin reuptake inhibitor; SNRI = serotonin-norepinephrine reuptake inhibitor.

### Treatment Efficacy

For each antidepressant selected, participants rated its overall effectiveness in reducing symptoms of depression as “Yes, a lot,” “Yes, a little,” or “No.” Sertraline had the highest proportion of strong positive responses, with 36.9% of participants rating it as “Yes, a lot” effective. This was followed by venlafaxine (36.0%), escitalopram (34.0%), citalopram (30.6%), and fluoxetine (28.6%). In contrast, tricyclic antidepressants had substantially lower rates, with only 14.2% of users overall reporting “Yes, a lot” effectiveness, including amitriptyline (12.7%), nortriptyline (14.3%), and imipramine (0%). Figure 3a displays the full distribution of effectiveness ratings for each antidepressant. When aggregated by drug class, SSRIs remained the most frequently prescribed and most rated as effective overall, followed by SNRIs, atypical antidepressants, and tricyclics (Figure 3b). Table S7 and S8 provide a detailed breakdown of all effectiveness ratings by individual antidepressant and drug class.

**Figure 3a.**
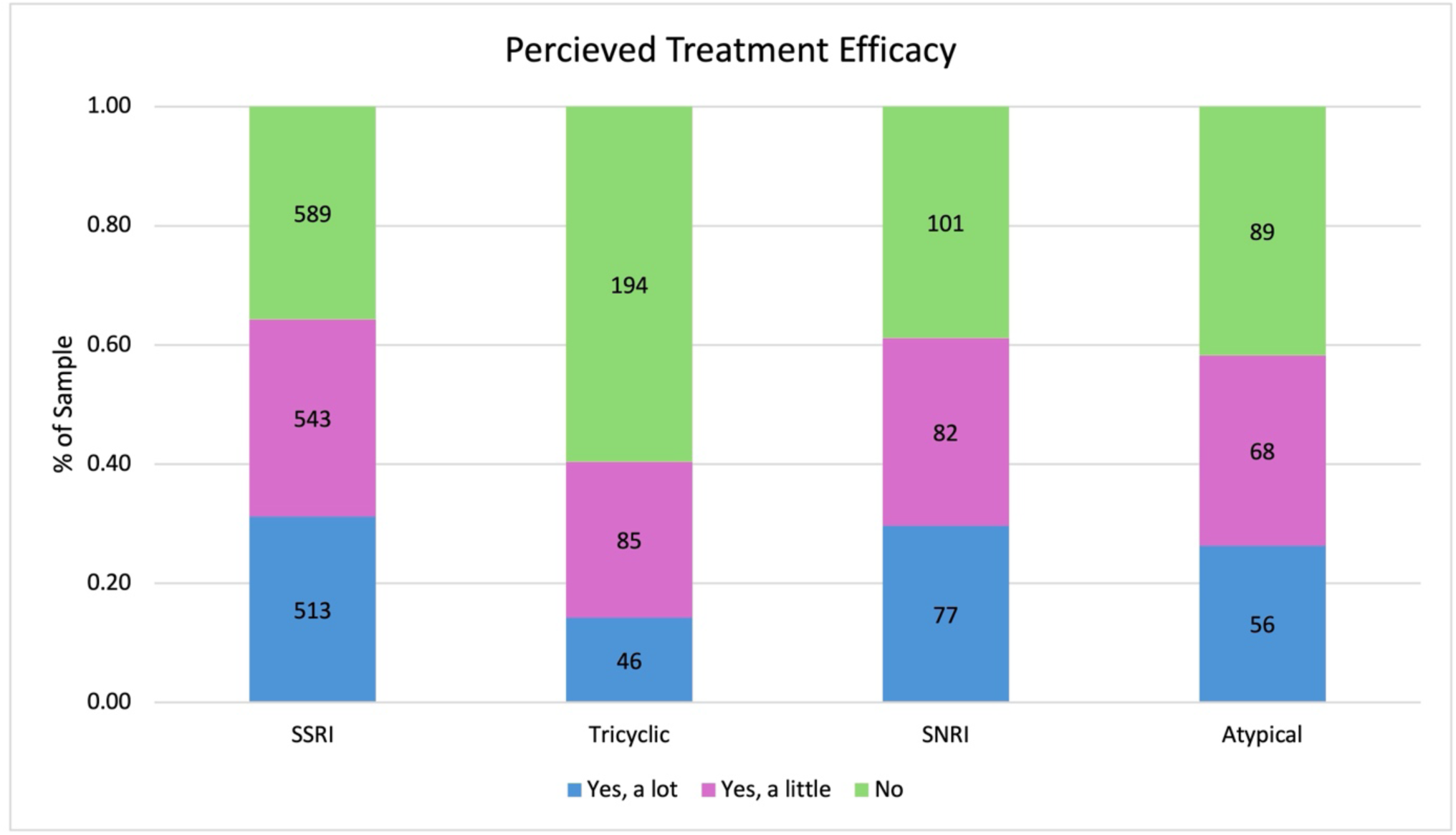
Perceived treatment efficacy of individual antidepressants. Stacked bar chart showing participant-reported effectiveness of individual antidepressants in reducing depressive symptoms. Participant responses to the question “Overall, do you feel [medication] is effective in reducing symptoms of depression?” are shown for each antidepressant. Responses are grouped by perceived effectiveness: “Yes, a lot” (blue), “Yes, a little” (pink), or “No” (green) effective. Numbers within bars indicate the count of participants in each response category for each antidepressant.

**Figure 3b.**
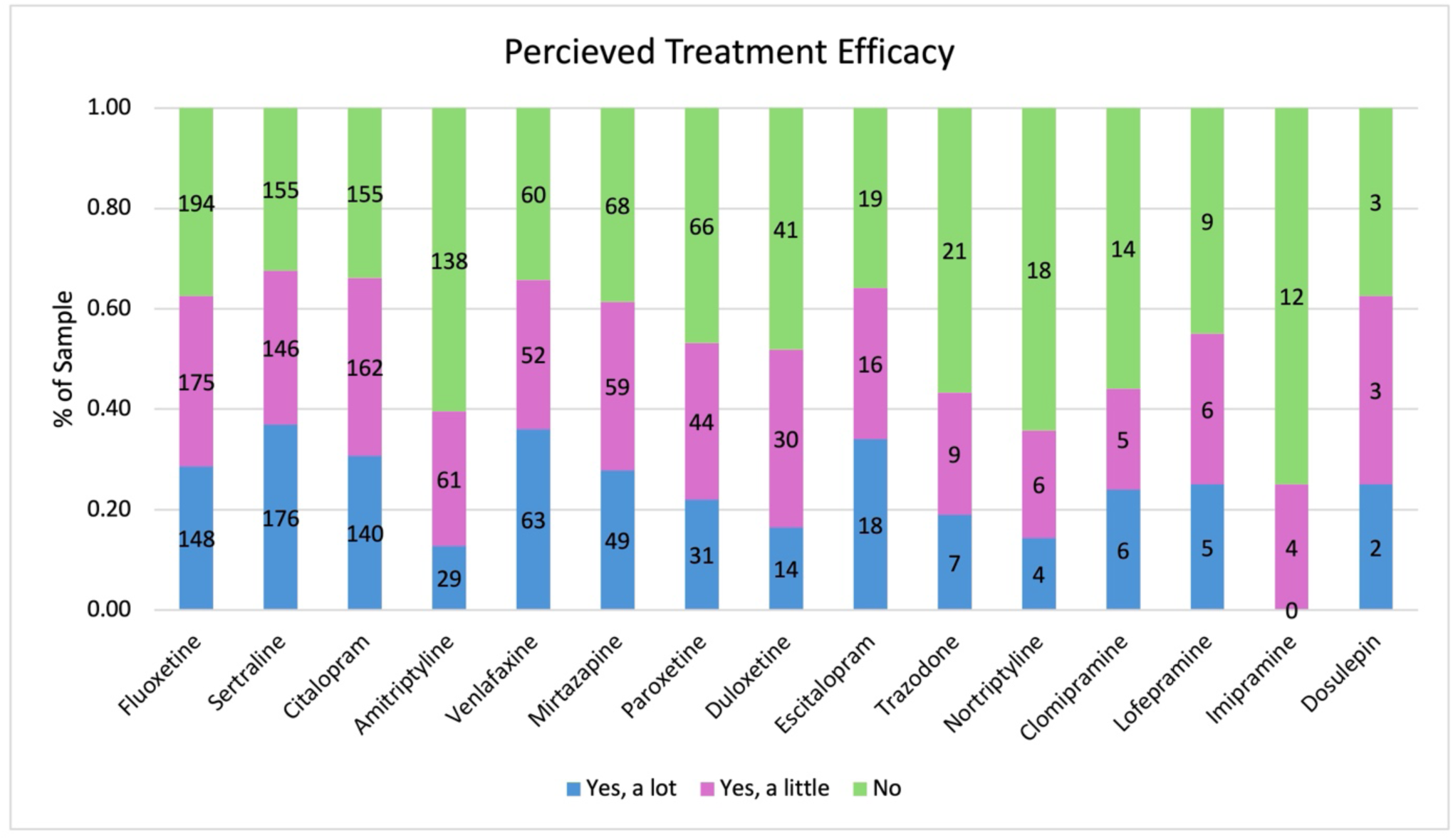
Perceived treatment efficacy aggregated by antidepressant class. Stacked bar chart showing participant-reported effectiveness aggregated by antidepressant class. Participant responses to the question “Overall, do you feel [medication] is effective in reducing symptoms of depression?” are grouped by drug class: SSRI (selective serotonin reuptake inhibitors), SNRI (serotonin-norepinephrine reuptake inhibitors), tricyclic, and atypical antidepressants. Responses are grouped by perceived effectiveness: “Yes, a lot” (blue), “Yes, a little” (pink), or “No” (green) effective. Numbers within bars indicate the count of participants in each response category for each antidepressant class.

Participants were asked to reflect on the specific symptom domains in which they noticed improvement from the antidepressant(s) they took. This was captured using the question: “Please rate the impact of the [medication] that helped in improving the following areas of symptoms that you have reported you experience when feeling depressed?” For each selected antidepressant, participants could rate its impact on the symptom domains: mood, anxiety, cognitive function, sleep, behaviour, and physical symptoms. Response options ranged from no impact (0) to extreme impact (6).

SSRIs had the highest mean impact on mood (2.12 on a 0-6 scale), anxiety (1.91), cognitive function (1.57), and behaviour (1.62) symptoms, and while atypical antidepressants had the highest impact on sleep symptoms (2.33), and SNRIs had the highest impact on physical symptoms (1.46). A summary of average symptom domain improvement aggregated by antidepressant class is presented in Table 4. Table S9 presents the mean impact ratings of all antidepressants on each symptom domain.

**Table 4.** Average reported improvement in symptom domains by an5depressant class. Improvement on a 7-point scale from no impact (0) to extreme impact (6). Mean improvement ratings across six symptom domains (Mood, Anxiety, Cognitive Function, Sleep, Behaviour, Physical) for each antidepressant class. Participants rated improvement on a 7-point scale from no impact (0) to extreme impact (6). SSRIs showed the highest mean improvement for mood (2.12), anxiety (1.91), cognitive function (1.57), and behaviour symptoms (1.62). Atypical antidepressants showed the highest improvement for sleep symptoms (2.33) and SNRIs showed the highest improvement for physical symptoms (1.34). SSRI = selective serotonin reuptake inhibitor; SNRI = serotonin-norepinephrine reuptake inhibitor; SD = standard deviation.

| Symptom Domain | Mean (SD) |  |  |  |
| --- | --- | --- | --- | --- |
|  | SSRI | Atypical | SNRI | Tricyclic |
| Mood | 2.12 (0.22) | 1.46 (0.38) | 1.88 (0.65) | 1.39 (0.72) |
| Anxiety | 1.91 (0.22) | 1.25 (0.43) | 1.64 (0.45) | 1.16 (0.66) |
| Cognitive Function | 1.57 (0.15) | 1.13 (0.23) | 1.38 (0.52) | 1.10 (0.52) |
| Sleep | 1.36 (0.09) | 2.33 (0.15) | 1.38 (0.16) | 1.79 (0.83) |
| Behaviour | 1.62 (0.21) | 1.27 (0.42) | 1.57 (0.47) | 1.40 (0.77) |
| Physical | 1.34 (0.13) | 1.02 (0.36) | 1.46 (0.22) | 1.27 (0.54) |

Participants were also asked to report the length of time it took for their symptoms to improve for each antidepressant they had taken. Time to improvement was rated on a scale from 0 (no improvement) to 6 (>6 months), with intermediate values representing 1-2 weeks (1), 2-4 weeks (2), 1-2 months (3), 2-4 months (4), and 4-6 months (5). Most antidepressants fell within the 2-3 range, corresponding to improvement between 2 to 4 weeks and 1 to 2 months. Table 5 describes the average time to improvement for each antidepressant.

**Table 5.** Participant reported average time to improvement for each antidepressant. The Improvement Category represents the mean 5me to ini5al symptom improvement, measured on a 7-point scale: 0 = no improvement no5ced, 1 = 1-2 weeks, 2 = 2-4 weeks, 3 = 1-2 months, 4 = 2-4 months, 5 = 4-6 months, and 6 = 6+ months. Number of participants who rated time to improvement, experienced improvement, or reported no improvement for each antidepressant. The Improvement Category represents the mean time to initial symptom improvement, measured on a 7-point scale: 0 = no improvement noticed, 1 = 1-2 weeks, 2 = 2-4 weeks, 3 = 1-2 months, 4 = 2-4 months, 5 = 4-6 months, and 6 = 6+ months. Most antidepressants showed mean improvement times between 2-4 weeks (category 2-3), with lofepramine showing the longest time to improvement (3.23) and trazodone the shortest (2.09). SD = standard deviation.

| Antidepressant | Rated time to improvement<br>N | Improved<br>N | No Improvement<br>N | Improvement Category<br>Mean (SD) |
| --- | --- | --- | --- | --- |
| Fluoxetine | 505 | 365 | 140 | 2.77 (1.08) |
| Sertraline | 474 | 367 | 107 | 2.9 (1.16) |
| Citalopram | 451 | 341 | 110 | 2.8 (1.16) |
| Amitriptyline | 233 | 153 | 80 | 2.41 (1.35) |
| Venlafaxine | 175 | 134 | 41 | 2.97 (1.19) |
| Mirtazapine | 176 | 133 | 43 | 2.67 (1.28) |
| Paroxetine | 131 | 93 | 38 | 2.59 (1.06) |
| Duloxetine | 86 | 58 | 28 | 2.93 (1.27) |
| Escitalopram | 49 | 39 | 10 | 2.62 (0.96) |
| Trazodone | 32 | 22 | 10 | 2.09 (1.11) |
| Nortriptyline | 28 | 15 | 13 | 3.07 (1.75) |
| Clomipramine | 27 | 15 | 12 | 2.53 (0.92) |
| Lofepramine | 17 | 13 | 4 | 3.23 (1.42) |
| Imipramine | 18 | 9 | 9 | 2.44 (1.74) |
| Dosulepin | 7 | 5 | 2 | 2.4 (0.55) |

### Side effects

The questionnaire captured self-reported side effects for each antidepressant the participants had taken. Participants were shown a predefined list of 20 common antidepressant side effects and could select any they had experienced while on each medication, along with “other” where they could write in a side effect. For this section, we omitted any free text answers. Across all antidepressants, the most reported side effects were sleep disturbances (insomnia or excessive sleepiness) (33.7% of participants), weight changes (29.7%), nausea (28.4%), fatigue or lack of energy (27.4%), and dry mouth (26.1%). Table 6 summarises the number of users who endorsed any side effect across all antidepressants.

**Table 6.** Participant reported prevalence of side effects. Number and percentage of participants endorsing each side effect across all antidepressants. Participants could report multiple side effects. Side effects are ordered by frequency of endorsement. The most commonly reported side effects were sleep disturbances (33.7%), weight gain or loss (29.7%), and nausea (28.4%).

| Side Effect | Prevalence<br>N | % of sample<br>endorsing |
| --- | --- | --- |
| Sleep disturbances (insomnia or excessive sleepiness) | 380 | 33.7 |
| Weight gain or loss | 334 | 29.7 |
| Nausea | 319 | 28.4 |
| Fatigue/lack of energy | 308 | 27.4 |
| Dry mouth | 294 | 26.1 |
| Sexual function or interest | 294 | 26.1 |
| Dizziness | 251 | 22.5 |
| Increased anxiety/agitation | 228 | 20.2 |
| Appetite changes | 226 | 20.1 |
| Attention/concentration difficulties | 205 | 18.2 |
| Headaches | 154 | 13.7 |
| Diarrhoea or constipation | 153 | 13.6 |
| Feeling/being sick | 151 | 13.4 |
| Memory problems | 139 | 12.3 |
| Shaking | 130 | 11.5 |
| Fast heartbeat | 122 | 10.3 |
| Indigestion or stomach aches | 119 | 10.1 |
| Muscle pain | 58 | 4.9 |
| Itching/rash | 53 | 4.5 |
| Blurred vision | 50 | 4.2 |

The antidepressant with the highest frequency of participants endorsing any side effects was venlafaxine with 43.7% of users endorsing at least one side effect, followed by mirtazapine (43.5%), and nortriptyline (43.3%) (Table 7). Detailed frequencies for individual side effects for all antidepressants and by drug class are provided in Tables S10-12.

**Table 7.** Proportion of participants that endorsed at least one side effect. Number of users for each antidepressant, number who endorsed at least one side effect, and percentage endorsing side effects. Venlafaxine (SNRI) and mirtazapine (atypical) had the highest rates of side effect endorsement (43.7% and 43.5% respectively), while dosulepin (tricyclic) had the lowest (20.0%). SSRI = selective serotonin reuptake inhibitor; SNRI = serotonin-norepinephrine reuptake inhibitor.

| Antidepressant | Class | Users<br>N | Endorsed Side<br>Effects<br>N | % of sample that<br>endorsed side<br>effects |
| --- | --- | --- | --- | --- |
| Fluoxetine | SSRI | 556 | 199 | 35.8 |
| Sertraline | SSRI | 506 | 206 | 40.7 |
| Citalopram | SSRI | 492 | 184 | 37.4 |
| Amitriptyline | Tricyclic | 272 | 104 | 38.2 |
| Venlafaxine | SNRI | 197 | 86 | 43.7 |
| Mirtazapine | Atypical | 191 | 83 | 43.5 |
| Paroxetine | SSRI | 159 | 63 | 39.6 |
| Duloxetine | SNRI | 97 | 27 | 27.8 |
| Escitalopram | SSRI | 56 | 22 | 39.3 |
| Trazodone | Atypical | 37 | 14 | 37.8 |
| Nortriptyline | Tricyclic | 30 | 13 | 43.3 |
| Clomipramine | Tricyclic | 28 | 8 | 28.6 |
| Lofepramine | Tricyclic | 23 | 9 | 39.1 |
| Imipramine | Tricyclic | 20 | 8 | 40.0 |
| Dosulepin | Tricyclic | 10 | 2 | 20.0 |

### Classification of Antidepressant Response

Using data collected in the questionnaire, we developed a rule-based framework to classify participants as antidepressant responders, non-responders, or unclassified (Figure 4). Responders were defined as participants who had taken at least one antidepressant for ≥ 6 weeks, had not discontinued the medication due to inefficacy, and rated the antidepressant as “Yes, a lot” when asked whether the medication was effective in reducing depressive symptoms. These individuals either continued taking the medication or had discontinued because they “started feeling better,” and they did not report “No” to efficacy for any antidepressant. Non-responders were those who had ≥2 failed antidepressant trials, where each medication was taken for an adequate duration (≥6 weeks), and who rated all adequately-trialled antidepressants as ineffective (“No” to overall efficacy). Discontinuations made solely because of side effects were not counted towards these criteria. This framework was informed by standard definitions of treatment-resistant depression (TRD), in which non-response typically requires at least two adequate antidepressant trials (European Medicines Agency, 2025). Participants who did not meet criteria for either group were classified as unclassified responders. Although the questionnaire captured multiple antidepressant classes, the classification focused on SSRIs. Responses to non-SSRI medications were incorporated only to restrict classification (e.g., reporting inefficacy to any antidepressant prevented classification as a responder).

**Figure 4.**
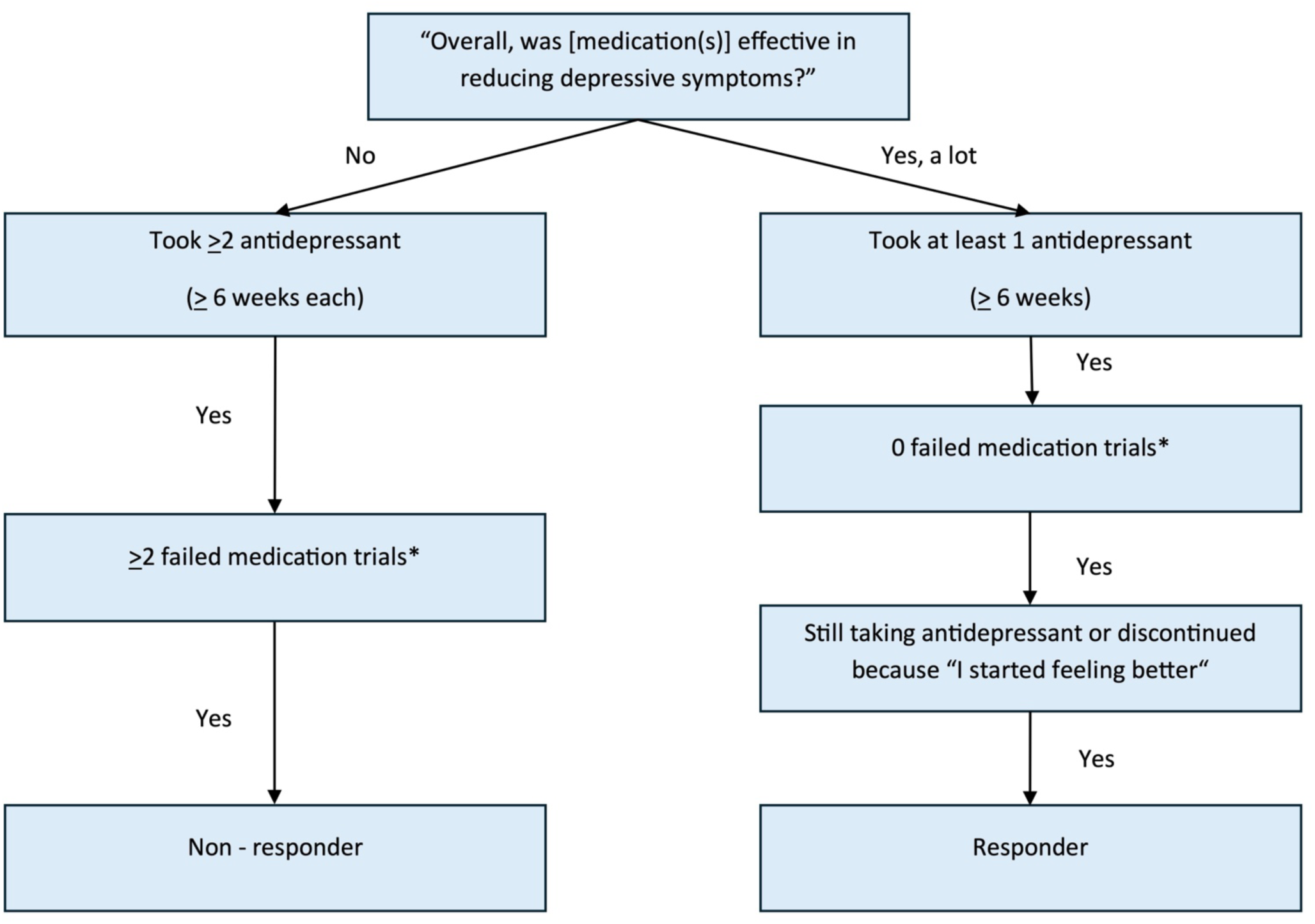
Rule-based framework for antidepressant response classification. Decision tree showing the classification criteria for identifying antidepressant responders and non-responders based on self-reported questionnaire data. Classification begins with participant response to the question “Overall, was [medication(s)] effective in reducing depressive symptoms?” Participants who answered “No” to all antidepressants and took ≥2 antidepressants for ≥6 weeks each were evaluated for non-responder classification. Non-responders were defined as participants with ≥2 failed medication trials, where each failed trial was an antidepressant taken for ≥6 weeks, discontinued due to inefficacy, and rated as ineffective. Participants who answered “Yes, a lot” for at least one antidepressant taken for ≥6 weeks were evaluated for responder classification. Responders were defined as participants with zero failed medication trials who were either still taking their effective antidepressant or discontinued because they started feeling better. Discontinuations due to side effects were not counted as failed trials. In this study, the framework was applied specifically to SSRIs, identifying 281 SSRI responders (23.8%) and 18 SSRI non-responders (1.5%), with 74.7% remaining unclassified. *Failed medication trials are defined as an antidepressant taken for ≥6 weeks, discontinued due to inefficacy, and rated as ineffective. Discontinuations due to side effects do not count as failed trials.

Applying these criteria, 281 participants (23.8%) were classified as SSRI responders, 18 participants (1.5%) as SSRI non-responders, and the remaining 881 (74.7%) as unclassified responders based on questionnaire data alone.

### Characteristics of Responders and Non-responders

Responders (n = 281) had lower baseline mean PHQ-9 (6.24) and GAD-7 (4.73) scores at the time of their original GS enrolment, compared with non-responders (n = 18) who had baseline mean PHQ-9 (13.11) and GAD-7 (11.89) scores, consistent with their classifications (Table S13). When comparing symptomology assessed within the questionnaire, lack of energy was the most endorsed symptom among responders (96.4%). All non-responders (n=18) endorsed lack of energy, racing thoughts, and depressed mood: sadness, hopelessness, or frequent crying. The top 20 endorsed symptoms for both groups are provided in Tables S14 and S15.

The average number of antidepressants taken for responders was 1.38 and 3.50 and for non-responders. Sertraline was the most used antidepressant across both responder (35.9%) and non-responder (83.3%) groups (Table S16). Among responders who had taken sertraline, 93.1% rated it as “yes, a lot” effective in reducing symptoms of depression, and the remaining 6.9% selected “yes, a little” (Table S17). Within our classification approach, non-responders are those who reported no antidepressant as effective. The most common side effect reported by responders was sexual dysfunction (40.4%), while for non-responders sexual dysfunction, dizziness, and sleep disturbances were equally common (each endorsed by 26.1%). Detailed medication usage, efficacy, and side effect frequencies for both groups are presented in Tables S16-S18.

### SNRI Responders

A small subgroup emerged among participants who initially met criteria for SSRI non-response but did not remain in the final non-responder category because they subsequently reported a positive response to an SNRI. Nine individuals fell into this category; six found venlafaxine effective and three found duloxetine effective. Across these participants, there were 24 SSRI trials, of which 23 (96%) were discontinued due to inefficacy and no medications were discontinued due to side effects. This pattern indicates that discontinuation was primarily driven by pharmacological non-response, rather than poor tolerability.

Symptomatically, all nine participants endorsed suicidal feelings or thoughts, depressed mood: sadness/hopelessness/frequent crying, guilt/self-blame, worthlessness and self-doubt, loss of interest in hobbies, avoidance of public areas, “everything feels like a burden”, reduced ability to make decisions, waking unrefreshed/always fatigued, reduced daytime activity, loss of interest in sex, lack of energy, and headaches/migraines (Table S20). The symptom with the highest impact on day-to-day life was avoidance of public areas with a mean (SD) impact of 4.56 (0.53). This profile reflects a severe, anergic (characterised by extreme fatigue and lack of energy), and somatic form of depression, contrasting with the anxiety/irritability and insomnia subtypes typically responsive to SSRIs (Lin & Stevens, 2014; Murawiec & Krzystanek, 2021). When compared with the broader SSRI responder group (n = 281), the greatest differences in prevalence were suicidal feelings or thoughts, headaches or migraines, oversleeping, and feeling detached from reality (Table 8), suggesting a distinct noradrenergic-related symptom phenotype. Tables S19-22 present symptom, antidepressant usage and efficacy, and side effect details for this group.

**Table 8.** Differences in symptom prevalence between SNRI responders and SSRI responders. Comparison of depression symptom endorsement rates between participants who responded to SNRIs (n=9) versus SSRIs (n=281). Symptoms shown are the top 10 with the largest differences in prevalence between groups, ordered by difference magnitude. Suicidal feelings or thoughts showed the largest difference between groups (61.2% difference), with 100% of SNRI responders endorsing this symptom compared to 38.8% of SSRI responders. SNRI = serotonin-norepinephrine reuptake inhibitor; SSRI = selective serotonin reuptake inhibitor.

| Domain | Symptom | SNRI Responder<br>Prevalence<br>N (%) | SSRI Responder<br>Prevalence<br>N (%) | Difference<br>% |
| --- | --- | --- | --- | --- |
| Depression | Suicidal feelings or thoughts | 9 (100) | 106 (38.8) | 61.2 |
| Physical | Headaches/migraines | 9 (100) | 137 (48.9) | 51.1 |
| Behaviour | Impulsive spending | 8 (88.9) | 108 (38.7) | 50.2 |
| Anxiety | Avoidance of public areas | 9 (100) | 147 (53.5) | 46.5 |
| Depression | Feeling detached from reality | 8 (88.9) | 123 (46.1) | 42.8 |
| Sleep | Oversleeping | 7 (77.8) | 99 (35.9) | 41.8 |
| Depression | No feelings of affection | 8 (88.9) | 134 (49.1) | 39.8 |
| Behaviour | Alienate friends/family | 8 (88.9) | 150 (54.2) | 34.7 |
| Sleep | Waking up much later | 7 (77.8) | 121 (43.8) | 34.0 |
| Anxiety | Feeling suspicious | 6 (66.7) | 99 (35.9) | 30.8 |

Although small in number, this group aligns with previous evidence that SNRIs achieve greater remission of somatic and low-energy symptoms than SSRIs. Others have argued that this is likely due to their dual serotonergic and noradrenergic action (Fagiolini et al., 2023; Zajecka & Albano, 2004). The identification of this same pattern in a population-based, self-reported dataset underscores the potential of self-report data. Identifying similar patterns within a population-based self-report cohort suggests potential value in symptom-cluster based prescribing, where antidepressant selection aligns with dominant symptom domains rather than a uniform first-line approach.

## Discussion

This paper presents the first descriptive results from the described questionnaire delivered to the Generation Scotland cohort, which was designed to collect detailed information on depressive symptoms, antidepressant exposure, and treatment experiences from people with lived experience of antidepressant use. These data represent a foundational component of AMBER’s broader programme, which aims to understand the biological mechanisms underlying antidepressant action and variability in antidepressant treatment outcomes to generate more precise and clinically meaningful phenotypes of antidepressant response and non-response.

The Generation Scotland cohort proved uniquely suited for investigating antidepressant treatment exposure and response due to its combination of genetic data, electronic health record linkage, longitudinal follow-up, and established recontact mechanisms. This infrastructure enabled recruitment of a large sample with antidepressant exposure, collection of detailed patient-reported treatment experiences, and ability to validate through linkage to EHRs. The extensive data available in Generation Scotland also provide opportunities for future studies examining patterns of treatment response, gene-environment interactions, and long-term outcomes. This established platform can support additional investigations of depression treatment, medication response phenotyping, and precision psychiatry research beyond the current study.

Between July and November 2025, 1,180 participants with a history of antidepressant use for depression completed the questionnaire. Respondents were predominantly female with the majority self-identifying as White, and generally older than the wider GS cohort and the Scottish population. However, prior research has shown this is a common trend in epidemiological study participation (Galea & Tracy, 2007). Geographic representation spanned all regions and deprivation quintiles, indicating wide national coverage.

Heterogeneous depressive symptom patterns found within the cohort highlight the multidimensional nature of depression experienced by antidepressant users in the community. Most participants endorsed lack of energy, waking up feeling unrefreshed/always fatigued, and reduced social activity and frequently reported as having a substantial impact on daily functioning. This pattern reinforces that depression is a heterogeneous disorder that can present with somatic, cognitive, and behavioural symptoms that are not always captured by standard tools used to diagnose depression. Standard clinical questionnaires used to diagnose depression, such as the PHQ-9, assess a limited subset of depressive symptoms, potentially overlooking dimensions relevant to treatment response (Fried & Nesse, 2015). In contrast, the questionnaire deliberately incorporated a wider range of depression symptoms across varying domains, including cognitive function, somatic, and behavioural symptoms. This expanded coverage of symptoms enabled the detection of symptom patterns that might be missed by standard screening tools. For example, the high prevalence of energy-related symptoms combined with domain-specific ratings of antidepressant impact facilitated the identification of a subgroup who later responded to SNRIs and showed distinct patterns of suicidality, anergic, and somatic symptoms.

About half of the participants took more than one antidepressant over time and about one third of our sample reported taking three or more antidepressants. This reflects the common clinical pattern where individuals try several medications before finding one that is effective or tolerable (Li et al., 2025). Such trial-and-error prescribing approaches place a significant burden on patients and services, prolongs time to remission, and highlights the need for approaches that can predict treatment response earlier in the care pathway. Patterns of antidepressant usage within this sample aligned with clinical prescribing guidelines. SSRIs were the most prevalent class, with 89.1% of the sample taking at least one SSRI, consistent with NICE prescribing guidance which recommends SSRIs as first-line pharmacological treatment for depression (National Institute for Health and Care Excellence (NICE), 2022). For individual antidepressant medications, fluoxetine, sertraline, and citalopram were the most reported. Participants also perceived SSRIs as the most effective overall for reducing symptoms of depression. SSRIs were found to be particularly impactful for mood, anxiety, and cognitive function symptom domains. SNRIs showed stronger self-reported benefit for somatic symptoms in this cohort, supporting evidence that SNRIs may offer more benefit to those with a more somatic presentation of depression (Perahia et al., 2009; Zajecka & Albano, 2004). Side effects were prevalent in this cohort, with sleep disturbances, weight changes, nausea, and fatigue/lack of energy among the most frequently experienced, reflecting well-known tolerability burdens associated with antidepressant use (Wang et al., 2018).

The Sequenced Treatment Alternatives to Relieve Depression study (STAR*D) is the largest and most influential clinical trial of antidepressant effectiveness and is widely used as a benchmark in antidepressant response research. In STAR*D, approximately one-third of participants achieved a clinically meaningful response to their first-line antidepressant (Gaynes et al., 2009). Among participants in the present cohort who reported taking only one antidepressant, just over half of these participants selected “Yes, a lot” when asked whether the medication had helped in reducing symptoms of depression. However, these figures are not directly comparable, as the STAR*D trial evaluated treatment response prospectively within a structured clinical trial, whereas our estimates are based on retrospective self-report within a naturalistic cohort with no limit on the time taken to achieve response.

Using self-reported antidepressant duration, discontinuation patterns, and perceived efficacy, we developed an initial rule-based classification of antidepressant treatment response. The classification focused specifically on SSRIs, as this drug class will be the target for subsequent biological sampling and molecular profiling. Approximately one-quarter met responder criteria, and a small group were classified as non-responders, with the remaining unclassified. This high unclassified proportion reflects the intentionally strict classification criteria designed to identify individuals at the extremes of treatment response. By applying these stringent criteria, we aimed to create well-defined phenotypic groups suitable for genetic and molecular investigations of treatment response, prioritising classification certainty over sample size. Future linkage with electronic health records will validate self-reported treatment histories, including diagnoses, medication maintenance patterns, and switching history to refine phenotype accuracy.

One notable insight was the identification of a small subgroup who did not benefit from multiple SSRI trials but subsequently reported a positive response to an SNRI. This group was characterised by a distinct symptom profile, as all participants in this group endorsed suicidal feelings or thoughts, compared to about one third of SSRI responders, representing the largest symptom prevalence difference between the groups. The SNRI responder group also universally endorsed headaches/migraines, loss of interest in hobbies, “everything feels like a burden,” waking unrefreshed/always fatigued, reduced daytime activity, and lack of energy. This profile characterised by suicidality, somatic, and low energy symptoms suggests a potential noradrenergic subtype that may warrant alternative first-line prescribing pathways. Although this subgroup is small and findings are preliminary, this pattern reflects real-world heterogeneity in antidepressant treatment response and highlights this study’s strength in detecting phenotype-specific treatment response signals that are unlikely to surface in EHR-only or trial-based designs.

Understanding how people experience antidepressants in real-world settings is essential for addressing the ongoing challenge of unpredictable treatment response. Clinical trials provide estimates of average efficacy, but they rarely capture the complexity of symptom patterns, switching histories, and side effects that shape lived experience. By collecting detailed self-reported data on depressive symptoms, antidepressant treatment experiences, and combining this with electronic health record and molecular data, this approach advances the field toward more nuanced and patient-centred antidepressant response phenotyping. These data lay the groundwork for developing predictive models of response, identifying subgroups who may benefit from specific drug classes, and ultimately reducing trial-and-error prescribing. In doing so, this work contributes an important step toward precision mental health care where treatments are guided by individual profiles.

### Future Plans

The next step in this project will link the questionnaire data to participants’ electronic health records (EHRs) to validate the self-reported antidepressant response classifications developed in this paper. EHR linkage will allow confirmation of prescription histories, treatment duration, switching patterns, and recorded diagnoses, thereby enhancing the clinical validity of the derived phenotypes.

After linkage, validated phenotyping methods will be applied to EHR data to identify antidepressant responders and non-responders. Antidepressant responders will be defined as individuals with antidepressant maintenance, indicated by stable prescriptions (consistent in strength, formulation, and dosage for ≥ 6 weeks) that meet one of the following criteria: (1) a single prescription episode lasting ≥ 182 days, (2) consecutive prescription episodes totalling ≥ 182 days, or (3) three or more consecutive prescription episodes. Non-responders will be identified using treatment-resistant depression (TRD) criteria, defined as individuals who have ≥ 2 antidepressant switches without achieving maintenance. All participants included in these definitions will be pre-selected as having ‘pure MDD’, requiring ≥ 2 MDD-related diagnostic codes in their GP or hospital records and no recorded diagnoses of drug abuse, psychosis, alcohol abuse, anxiety disorders, bipolar disorder, or sleep disorders. Only antidepressant prescription episodes lasting ≥ 6 weeks will be included in the analysis.

We will then initiate biological sample collection, targeting 25 individuals classified as responders and 25 as non-responders, based on combined evidence from EHR and questionnaire data within the GS cohort. As questionnaire recruitment remains open and we begin EHR linkage, we anticipate identifying sufficient non-responders to meet sampling targets. If needed, classification criteria may be relaxed (e.g., requiring one failed SSRI trial rather than two) to ensure adequate sample size for molecular analyses. Each participant will provide both saliva and blood samples. Saliva and blood samples will undergo DNA extraction followed by methylation analysis. Blood samples will also be processed for peripheral blood mononuclear cell (PBMC) isolation. PBMCs will be used for downstream cellular assays, and the remaining PBMCs will be cryopreserved for future induced pluripotent stem cell (iPSC) work. A subset of PBMCs will be transformed into lymphoblastoid cell lines (LCLs), providing cellular material for gene expression assays. These cell lines will allow direct comparison of gene expression signatures between responders and non-responders, supporting further investigation of whether molecular pathways identified in commercial cell line screens generalise to patient-derived tissue. LCLs and PBMC-derived cultures will be exposed to SSRIs to generate antidepressant-induced gene expression signatures. Differential expression and pathway analyses will be used to identify cellular mechanisms and biological pathways that distinguish treatment response profiles. These molecular data will integrate with genomic, methylation, clinical, and EHR-derived information across the wider AMBER project to advance understanding of the biological mechanisms underpinning antidepressant response and non-response. Upon completion of the planned data integration, this work will establish a valuable resource for future antidepressant exposure and response research beyond the AMBER project. The combination of detailed patient-reported symptom profiles and medication histories with electronic health record linkage and molecular profiling will provide a uniquely comprehensive characterisation of antidepressant treatment response in a population-based sample.

### What are the main strengths and weaknesses?

The present study has several notable strengths. It leverages a large, population-based cohort embedded within GS, providing extensive demographic, genetic, and health-related data alongside new self-reported information from the questionnaire. The ability to link questionnaire responses to existing genetic, lifestyle, and NHS electronic health records offers substantial potential for longitudinal and multi-modal analyses of antidepressant use and treatment response. Furthermore, the questionnaire was co-developed with the Lived Experience Advisory Panel, ensuring that questions were clear, sensitive, and reflective of real-world treatment experiences, strengthening the validity and acceptability of the instrument. The questionnaire captures rich, participant-reported data on symptom experiences, medication histories, and perceived efficacy, offering valuable insight into the lived experience of antidepressant treatment at a population scale.

Several limitations should also be acknowledged. The demographic composition of the cohort, being primarily older (median age 57 years), female (78.1%), and self-identifying as White (98.6%), may limit the generalisability of these results to the full range of patients seen in clinical practice for depression and antidepressant treatment or to more diverse populations. However, this cohort still provides rich, detailed patient-reported data that captures symptom heterogeneity and treatment experiences not available in electronic health records alone and establishes a foundational dataset for developing and validating antidepressant response phenotypes. The methods and phenotyping framework developed here can be applied to more diverse populations in future studies.

The questionnaire data are self-reported and, therefore, subject to recall bias and misclassification of treatment histories and perceived efficacy. This is a common limitation of retrospective survey research (Althubaiti, 2016). We attempted to mitigate this through clear question wording developed with input from the Lived Experience Advisory Panel and by incorporating questions that focused on concrete experiences (e.g., medication names, duration, reasons for discontinuation) rather than requiring recall of dates or clinical details.

The questionnaire captured medication history, duration, discontinuation reasons, and efficacy for each antidepressant, but did not collect information on the timing between medications or whether multiple antidepressants were used within the same depressive episode versus across different episodes over time. Future linkage with electronic health records will provide prescription timelines to help clarify medication sequences and episode-specific treatment patterns.

The antidepressant response classification developed here is based on self-report and may not capture the full clinical complexity of treatment response, adherence, or comorbidity. However, these self-reported classifications will be validated and refined through future linkage with electronic health records and prescription data, which will provide objective verification of diagnoses, medication continuity, and switching behaviour, strengthening the clinical validity of the derived phenotypes.

### Data Availability

### Underlying data

The questionnaire data analysed in this paper were collected through the GS cohort. Access to data is governed by the GS Access Committee and is available to researchers upon application (https://genscot.ed.ac.uk/). Due to ethical and data protection restrictions, individual-level questionnaire responses and linked health data are not publicly available.

To learn more about the AMBER study, you can visit the study website here: https://www.kcl.ac.uk/research/amber-antidepressant-medications-biology-exposure-response.

### Source Code

Source code available from: https://github.com/megcal8/AMBER-Generation-Scotland

Archived source code available from: https://doi.org/10.5281/zenodo.18741590 (Edmondson-Stait et al., 2026)

License: MIT License

## Extended Data

Zenodo: Supplementary Material for Cohort Profile: Investigating Antidepressant Response within Generation Scotland. https://doi.org/10.5281/zenodo.18612355 (Calnan et al., 2026)

This project contains the following extended data:

- Supplementary Material

`o` Figure S1
`o` Tables S1-S22
- Questionnaire instrument adapted from AGDS:Cell-o

Data are available under the terms of the Creative Commons Zero “No rights reserved” data waiver (CC0 1.0 Public domain dedication).

## Ethical approval

Ethical approval was received from the West Midlands - South Birmingham Research Ethics Committee (REC) (REC reference: 24/WM/0264). Generation Scotland holds Research Tissue Bank REC approval from the East of Scotland Research Ethics Service (EoSRES) (REC reference: 25/ES/0013).

## Consent

Written informed consent for publication of the participants anonymised data details was obtained from the participants.

## Supporting information

Questionnaire

## Acknowledgements

We acknowledge the invaluable contributions of the other investigators on the AMBER: Antidepressant Medications: Biology, Exposure & Response project: Iona Beange, Cristina Douglas, Sue Fletcher-Watson, Mark Somerville, Emer O’Leary, Arlene Casey, Matús Falis, Mark J. Adams, Matthew H. Iveson, Emily L. Ball, Heather C. Whalley, Chris Wai Hang Lo, Ke Hu, Kate Stewart, Cathryn M. Lewis, Michelle Kamp, Oliver Pain, David Brici, Anjali K. Henders, Quan Nguyen, Sonia Shah, Naomi R. Wray, Clara Albiñana, and Alicia Walker.

We also express our appreciation to the members of AMBER’s Lived Experience Advisory Panel (LEAP), whose insights and feedback are integral to the design and delivery of the entire AMBER project. Finally, we are extremely grateful to all the people who are part of Generation Scotland and to the Generation Scotland team for their continued support and collaboration.

