## Supplementary Materials for "Cohort Profile: Investigating Antidepressant Response within Generation Scotland"

### Supplementary Material

Megan Calnan[1], Amelia Edmondson – Stait[1], Hannah Milbourn[2], Esme Elsden[1], Anjali K. Henders[4], Emily L. Ball[1], Matthew H. Iveson[1], AMBER Research Team, AMBER Lived Experience Advisory Panel, Generation Scotland Team, Naomi R. Wray[3,4], Sonia Shah[4], Cathryn M. Lewis[5,6], and Andrew M. McIntosh[1]

1. Institute for Neuroscience and Cardiovascular Research, University of Edinburgh, Edinburgh, United Kingdom
2. Centre for Genomic and Experimental Medicine, Institute of Genetics and Cancer, University of Edinburgh, Edinburgh, United Kingdom
3. Department of Psychiatry, University of Oxford, United Kingdom
4. Institute for Molecular Bioscience, University of Queensland, Brisbane, Queensland, Australia
5. Social, Genetic & Developmental Psychiatry Centre, King's College London, London, United Kingdom
6. Department of Medical and Molecular Genetics, King's College London, London, United Kingdom

**Contents:**

1. Supplementary Figures S1
2. Supplementary Tables S1-S22

**Figure S1.** Depression symptom domains and corresponding symptom items assessed in the questionnaire.

**Mood**
- Depressed Mood; sadness, hopelessness, frequent crying
- Feelings of Guilt: blaming yourself and feel you are failing
- Feelings of worthlessness and self-doubt
- Loss of interest in hobbies and in things you usually enjoyed
- I am feeling detached from reality
- Don't have feelings of affection even when you know you should
- Surroundings feel unreal or like looking through a thick fog
- Suicidal feelings or thinking about suicide

**Anxiety**
- Anxious – worrying about minor things
- Repetitive fearful thoughts - like being afraid of catching an infection, or that a disaster might happen
- Feeling suspicious of other people
- Intrusive, recurrent thoughts about your own health
- Engagement in very repetitive behaviours that are linked to your anxiety - like unnecessary washing, cleaning, or checking things over
- Panic attacks
- Avoidance of public areas going out in public or travelling outside your home
- Persistent focus on past traumatic events

**Cognitive Function**
- Reduced ability to think and process thoughts
- Everything feels like it is a burden: can't be bothered with anything
- Reduced ability to concentrate on a book or movie
- Reduced ability to concentrate at work/study
- Reduced ability to complete everyday tasks
- Reduced ability to manage the workload expected of you
- Racing thoughts - Your mind isn't able to "shut off" and you can't fully relax
- Reduced ability to make decisions
- Decreased creativity
- Increased ability to focus and manage the workload expected of you

**Sleep**
- Difficulty getting to sleep
- Waking throughout the night
- Waking up much later
- Oversleeping (sleep more than 8-10 hours a night)
- Waking up feeling unrefreshed, always feeling fatigued
- Feel worst in the morning
- Internal body clock seems out of sync with actual time of day - like jetlag
- Early morning (before sunrise) wakening

**Behaviour**
- Unable to sit still, feeling the need to keep moving
- Reduced daytime activity
- Increased sensitivity to criticism or rejection
- Spend money without thinking it through
- Easily irritated or frustrated - even unprovoked rage
- Alienate friends and/or family
- Reduced interest in participating in social activities
- Loss of interest in sex
- Engage very repetitive behaviours - like unnecessary washing, cleaning or checking over and over
- Reluctance to talk give single word answers when asked a question

**Physical**
- Lack of energy
- Everything is in slow motion
- Racing heart, sweat or have trouble breathing
- Appetite increases
- Appetite decreases
- Significant weight gain
- Significant weight loss
- Heavy feeling in arms or legs
- Unexplained aches and pains
- Headaches and/or migraines
- Gut problems e.g., constipation, peptic ulcers, reflux, irritable bowel
- Low sex drive
- Sexual dysfunction
- Changes in menstrual cycle

**Table S1**. Endorsement rates and mean impact ratings of symptoms on daily function within the mood domain. Impact rated on a 5-point scale: 1 (no impact) to 5 (severe impact).

| Symptom | Endorsed N (%) | Impact Mean (SD) |
|---|---|---|
| Loss of interest in hobbies | 1028 (88.6) | 3.44 (0.92) |
| Depressed mood: sadness, hopelessness, frequent crying | 1018 (87) | 3.5 (0.86) |
| Worthlessness and self-doubt | 974 (83.6) | 3.54 (0.91) |
| Guilt: blaming yourself | 891 (77.1) | 3.38 (0.91) |
| No feelings of affection | 650 (56.3) | 3.4 (0.99) |
| Feeling detached from reality | 596 (51.9) | 3.15 (1.01) |
| Suicidal feelings or thoughts | 559 (48.4) | 3.35 (1.13) |
| Surroundings feel unreal/foggy | 476 (41.3) | 3.3 (0.95) |

**Table S2.** Endorsement rates and mean impact ratings of symptoms on daily function within the anxiety domain. Impact rated on a 5-point scale: 1 (no impact) to 5 (severe impact).

| Symptom | Endorsed N (%) | Impact Mean (SD) |
|---|---|---|
| Anxious/worrying | 1004 (85.2) | 3.41 (0.93) |
| Avoidance of public areas | 746 (63.9) | 3.68 (0.98) |
| Persistent focus on past trauma | 691 (59.2) | 3.51 (1.02) |
| Intrusive thoughts about health | 641 (55.1) | 3.27 (0.92) |
| Repetitive fearful thoughts | 530 (45.3) | 3.68 (0.87) |
| Panic attacks | 526 (45) | 3.51 (0.98) |
| Feeling suspicious | 487 (41.8) | 3.2 (1.01) |
| Repetitive behaviours | 308 (26.4) | 3.33 (0.95) |

**Table S3.** Endorsement rates and mean impact ratings of symptoms on daily function within the cognitive function domain. Impact rated on a 5-point scale: 1 (no impact) to 5 (severe impact).

| Symptom | Endorsed N (%) | Impact Mean (SD) |
|---|---|---|
| Everything feels like a burden | 1028 (87.4) | 3.6 (0.91) |
| Racing thoughts | 1010 (86) | 3.69 (0.97) |
| Can't concentrate on book/movie | 983 (83.7) | 3.12 (0.96) |
| Reduced ability to think | 942 (80.4) | 3.44 (0.89) |
| Can't concentrate at work/study | 936 (79.7) | 3.48 (0.93) |
| Decreased creativity | 915 (78.5) | 2.93 (1) |
| Reduced ability to make decisions | 907 (77.4) | 3.46 (0.9) |
| Can't complete everyday tasks | 898 (76.5) | 3.52 (0.92) |
| Can't manage workload | 853 (73.1) | 3.5 (0.93) |
| Increased focus/workload ability | 403 (34.5) | 3.32 (1.04) |

**Table S4.** Endorsement rates and mean impact ratings of symptoms on daily function within the sleep domain. Impact rated on a 5-point scale: 1 (no impact) to 5 (severe impact).

| Symptom | Endorsed N (%) | Impact Mean (SD) |
|---|---|---|
| Waking unrefreshed/always fatigued | 1096 (93) | 3.78 (0.9) |
| Waking throughout night | 966 (82.1) | 3.48 (0.91) |
| Difficulty getting to sleep | 908 (77.1) | 3.66 (0.9) |
| Feel worst in morning | 802 (68.4) | 3.46 (0.96) |
| Early morning wakening | 716 (61) | 3.23 (1.02) |
| Body clock out of sync | 659 (56.2) | 3.44 (0.94) |
| Waking up much later | 520 (44.6) | 3.31 (0.97) |
| Oversleeping | 426 (36.5) | 3.46 (1.07) |

**Table S5.** Endorsement rates and mean impact ratings of symptoms on daily function within the behaviour domain. Impact rated on a 5-point scale: 1 (no impact) to 5 (severe impact).

| Symptom | Endorsed N (%) | Impact Mean (SD) |
|---|---|---|
| Reduced social activity | 1068 (90.8) | 3.34 (0.95) |
| Sensitive to criticism or rejection | 1038 (88.3) | 3.5 (0.98) |
| Reduced daytime activity | 911 (77.6) | 3.29 (0.87) |
| Loss of interest in sex | 895 (77.4) | 2.99 (1.24) |
| Easily irritated | 847 (72.1) | 3.54 (0.97) |
| Reluctance to talk | 726 (61.8) | 3.25 (0.96) |
| Alienate friends/family | 724 (61.8) | 3.53 (0.96) |
| Impulsive spending | 561 (47.9) | 3.54 (1.02) |
| Restless/need to keep moving | 375 (31.9) | 3.12 (0.97) |
| Repetitive behaviours | 270 (23) | 3.25 (1.03) |

**Table S6.** Endorsement rates and mean impact ratings of symptoms on daily function within the physical domain. Impact rated on a 5-point scale: 1 (no impact) to 5 (severe impact).

| Symptom | Endorsed N (%) | Impact Mean (SD) |
|---|---|---|
| Lack of energy | 1121 (95.1) | 3.66 (0.93) |
| Low sex drive | 835 (72.4) | 2.79 (1.2) |
| Gut problems | 718 (61.4) | 3.57 (0.92) |
| Unexplained aches and pains | 663 (56.6) | 3.29 (0.91) |
| Headaches/migraines | 622 (52.8) | 3.37 (0.93) |
| Significant weight gain | 555 (47.3) | 3.71 (0.92) |
| Appetite increases | 551 (47.1) | 3.4 (0.97) |
| Racing heart/sweating | 522 (44.5) | 3.38 (0.99) |
| Heavy feeling in limbs | 498 (42.6) | 3.06 (0.91) |
| Appetite decreases | 492 (42.3) | 2.79 (1.02) |
| Everything in slow motion | 445 (38) | 3.14 (0.97) |

| | | |
|---|---|---|
| Sexual dysfunction | 434 (37.9) | 3.04 (1.2) |
| Significant weight loss | 173 (15) | 2.97 (1.09) |
| Changes in menstrual cycle | 112 (11.8) | 3.27 (0.99) |

**Table S7.** Perceived treatment efficacy of individual antidepressants. Participants reported whether the medication was effective in reducing symptoms of depression ("Yes, a lot," "Yes, a little," or "No").

| Antidepressant | Class | Rated efficacy<br>Rated efficacy<br>N | Yes, a lot<br>N (%) | Yes, a little<br>N (%) | No<br>N (%) |
|---|---|---|---|---|---|
| Fluoxetine | SSRI | 517 | 148 (28.6) | 175 (33.8) | 194 (37.5) |
| Sertraline | SSRI | 477 | 176 (36.9) | 146 (30.6) | 155 (32.5) |
| Citalopram | SSRI | 457 | 140 (30.6) | 162 (35.4) | 155 (33.9) |
| Amitriptyline | Tricyclic | 228 | 29 (12.7) | 61 (26.8) | 138 (60.5) |
| Venlafaxine | SNRI | 175 | 63 (36.0) | 52 (29.7) | 60 (34.3) |
| Mirtazapine | Atypical | 176 | 49 (27.8) | 59 (33.5) | 68 (38.6) |
| Paroxetine | SSRI | 141 | 31 (22.0) | 44 (31.2) | 66 (46.8) |
| Duloxetine | SNRI | 85 | 14 (16.5) | 30 (35.3) | 41 (48.2) |
| Escitalopram | SSRI | 53 | 18 (34.0) | 16 (30.2) | 19 (35.8) |
| Trazodone | Atypical | 37 | 7 (18.9) | 9 (24.3) | 21 (56.8) |
| Nortriptyline | Tricyclic | 28 | 4 (14.3) | 6 (21.4) | 18 (64.3) |
| Clomipramine | Tricyclic | 25 | 6 (24.0) | 5 (20.0) | 14 (56.0) |
| Lofepramine | Tricyclic | 20 | 5 (25.0) | 6 (30.0) | 9 (45.0) |
| Imipramine | Tricyclic | 16 | 0 (0.0) | 4 (25.0) | 12 (75.0) |
| Dosulepin | Tricyclic | 8 | 2 (25.0) | 3 (37.5) | 3 (37.5) |

**Table S8.** Perceived treatment efficacy aggregated by antidepressant class.

| Antidepressant Class | Rated efficacy<br>N | Yes, a lot<br>N (%) | Yes, a little<br>N (%) | No<br>N (%) |
|---|---|---|---|---|
| SSRI | 1645 | 513 (31.2) | 543 (33.0) | 589 (35.8) |
| Tricyclic | 325 | 46 (14.2) | 85 (26.2) | 194 (59.7) |
| SNRI | 260 | 77 (29.6) | 82 (31.5) | 101 (38.8) |
| Atypical | 213 | 56 (26.3) | 68 (31.9) | 89 (41.8) |

**Table S9.** Mean reported improvement by symptom domain for each antidepressant. Impact rated on a 7-point scale: no impact (0) to extreme impact (6).

| Antidepressant | Antidepressant Class | Domain | Rated Improvement<br>N | Impact<br>Mean (SD) |
|---|---|---|---|---|
| Fluoxetine | SSRI | Mood | 527 | 2.12 (1.75) |
| | | Anxiety | 525 | 1.75 (1.7) |
| | | Cognitive Function | 526 | 1.56 (1.59) |
| | | Sleep | 528 | 1.34 (1.48) |
| | | Behaviour | 528 | 1.7 (1.6) |

| | | | | |
|---|---|---|---|---|
| | | Physical | 526 | 1.28 (1.38) |
| Sertraline | SSRI | Mood | 484 | 2.39 (1.79) |
| | | Anxiety | 483 | 2.13 (1.76) |
| | | Cognitive Function | 480 | 1.76 (1.61) |
| | | Sleep | 482 | 1.51 (1.57) |
| | | Behaviour | 481 | 1.92 (1.68) |
| | | Physical | 482 | 1.53 (1.57) |
| Citalopram | SSRI | Mood | 470 | 2.17 (1.73) |
| | | Anxiety | 470 | 1.99 (1.76) |
| | | Cognitive Function | 460 | 1.58 (1.57) |
| | | Sleep | 475 | 1.37 (1.48) |
| | | Behaviour | 465 | 1.65 (1.56) |
| | | Physical | 467 | 1.4 (1.48) |
| Amitriptyline | Tricyclic | Mood | 256 | 1.14 (1.43) |
| | | Anxiety | 255 | 1.16 (1.46) |
| | | Cognitive Function | 254 | 0.98 (1.39) |
| | | Sleep | 256 | 2.19 (1.94) |
| | | Behaviour | 254 | 1 (1.38) |
| | | Physical | 254 | 1.19 (1.52) |
| Venlafaxine | SNRI | Mood | 186 | 2.34 (1.84) |
| | | Anxiety | 186 | 1.95 (1.81) |
| | | Cognitive Function | 185 | 1.75 (1.65) |
| | | Sleep | 185 | 1.49 (1.64) |
| | | Behaviour | 186 | 1.9 (1.67) |
| | | Physical | 186 | 1.61 (1.58) |
| Mirtazapine | Atypical | Mood | 181 | 1.73 (1.73) |
| | | Anxiety | 181 | 1.56 (1.69) |
| | | Cognitive Function | 180 | 1.29 (1.52) |
| | | Sleep | 182 | 2.43 (2.01) |
| | | Behaviour | 180 | 1.56 (1.54) |
| | | Physical | 179 | 1.27 (1.57) |
| Paroxetine | SSRI | Mood | 142 | 1.78 (1.67) |
| | | Anxiety | 143 | 1.61 (1.57) |
| | | Cognitive Function | 141 | 1.34 (1.44) |
| | | Sleep | 142 | 1.27 (1.43) |
| | | Behaviour | 144 | 1.43 (1.51) |
| | | Physical | 142 | 1.27 (1.48) |
| Duloxetine | SNRI | Mood | 91 | 1.42 (1.74) |
| | | Anxiety | 91 | 1.32 (1.7) |
| | | Cognitive Function | 91 | 1.02 (1.41) |
| | | Sleep | 90 | 1.26 (1.59) |
| | | Behaviour | 90 | 1.24 (1.57) |

| | | | | |
|---|---|---|---|---|
| | | Physical | 91 | 1.3 (1.66) |
| Escitalopram | SSRI | Mood | 50 | 2.14 (1.81) |
| | | Anxiety | 50 | 2.06 (1.73) |
| | | Cognitive Function | 51 | 1.59 (1.7) |
| | | Sleep | 51 | 1.31 (1.45) |
| | | Behaviour | 51 | 1.41 (1.54) |
| | | Physical | 51 | 1.2 (1.54) |
| Trazodone | Atypical | Mood | 37 | 1.19 (1.54) |
| | | Anxiety | 37 | 0.95 (1.39) |
| | | Cognitive Function | 37 | 0.97 (1.4) |
| | | Sleep | 37 | 2.22 (1.93) |
| | | Behaviour | 36 | 0.97 (1.4) |
| | | Physical | 37 | 0.76 (1.34) |
| Nortriptyline | Tricyclic | Mood | 27 | 0.48 (0.94) |
| | | Anxiety | 28 | 0.36 (0.87) |
| | | Cognitive Function | 28 | 0.36 (1.1) |
| | | Sleep | 28 | 1.04 (1.35) |
| | | Behaviour | 28 | 0.5 (1.07) |
| | | Physical | 28 | 0.68 (1.16) |
| Clomipramine | Tricyclic | Mood | 26 | 1.65 (1.72) |
| | | Cognitive Function | 26 | 1.5 (1.75) |
| | | Sleep | 26 | 1.35 (1.67) |
| | | Behaviour | 26 | 1.5 (1.63) |
| | | Physical | 26 | 1.27 (1.43) |
| Lofepramine | Tricyclic | Mood | 21 | 1.57 (1.43) |
| | | Anxiety | 21 | 1.33 (1.49) |
| | | Cognitive Function | 20 | 1 (1.26) |
| | | Sleep | 21 | 1.57 (1.47) |
| | | Behaviour | 20 | 1.45 (1.43) |
| | | Physical | 20 | 1.15 (1.39) |
| Imipramine | Tricyclic | Mood | 16 | 0.94 (1.24) |
| | | Anxiety | 16 | 0.81 (1.17) |
| | | Cognitive Function | 16 | 0.88 (1.15) |
| | | Sleep | 16 | 1.31 (1.85) |
| | | Behaviour | 16 | 1 (1.46) |
| | | Physical | 16 | 1.06 (1.53) |
| Dosulepin | Tricyclic | Mood | 7 | 2.57 (1.99) |
| | | Anxiety | 7 | 2.14 (2.48) |
| | | Cognitive Function | 7 | 1.86 (2.12) |
| | | Sleep | 7 | 3.29 (2.14) |
| | | Behaviour | 7 | 2.57 (1.99) |
| | | Physical | 7 | 2.29 (1.89) |

**Table S10**. Prevalence of side effects for each antidepressant.

| Side Effect | Prevalence N (%) | | | | | | | | | | | | | | |
|---|---|---|---|---|---|---|---|---|---|---|---|---|---|---|---|
| | Fluoxetine | Sertraline | Citalopram | Amitriptyline | Venlafaxine | Mirtazapine | Paroxetine | Duloxetine | Escitalopram | Trazodone | Nortriptyline | Clomipramine | Lofepramine | Imipramine | Dosulepin |
| Sleep disturbances | 74 (33.0) | 63 (28.6) | 59 (29.2) | 53 (47.3) | 31 (32.3) | 32 (37.2) | 19 (27.1) | 14 (42.4) | 8 (34.8) | 9 (56.2) | 7 (46.7) | 2 (22.2) | 4 (40.0) | 5 (62.5) | 0 (0.0) |
| Weight gain/loss | 54 (24.1) | 59 (26.8) | 64 (31.7) | 28 (25.0) | 28 (29.2) | 52 (60.5) | 18 (25.7) | 6 (18.2) | 8 (34.8) | 6 (37.5) | 5 (33.3) | 2 (22.2) | 3 (30.0) | 1 (12.5) | 0 (0.0) |
| Nausea | 53 (23.8) | 70 (31.8) | 70 (34.7) | 25 (22.3) | 30 (31.2) | 15 (17.4) | 24 (34.3) | 12 (36.4) | 6 (26.1) | 2 (12.5) | 4 (26.7) | 4 (44.4) | 1 (10.0) | 3 (37.5) | 0 (0.0) |
| Fatigue/lack of energy | 50 (22.3) | 59 (26.8) | 44 (21.8) | 45 (40.2) | 24 (25.0) | 25 (29.1) | 21 (30.0) | 8 (24.2) | 9 (39.1) | 8 (50.0) | 7 (46.7) | 2 (22.2) | 2 (20.0) | 3 (37.5) | 1 (50.0) |
| Dry mouth | 41 (18.3) | 54 (24.5) | 56 (27.7) | 38 (33.9) | 32 (33.3) | 17 (19.8) | 14 (20.0) | 8 (24.2) | 3 (13.0) | 2 (12.5) | 9 (60.0) | 3 (33.3) | 8 (80.0) | 7 (87.5) | 2 (100.0) |
| Sexual function or interest | 61 (27.2) | 79 (35.9) | 61 (30.2) | 8 (7.1) | 21 (21.9) | 14 (16.3) | 22 (31.4) | 9 (27.3) | 13 (56.5) | 3 (18.8) | 2 (13.3) | 0 (0.0) | 1 (10.0) | 0 (0.0) | 0 (0.0) |
| Dizziness | 29 (12.9) | 55 (25.0) | 48 (23.8) | 28 (25.0) | 26 (27.1) | 16 (18.6) | 18 (25.7) | 11 (33.3) | 5 (21.7) | 3 (18.8) | 6 (40.0) | 2 (22.2) | 1 (10.0) | 3 (37.5) | 0 (0.0) |
| Increased anxiety/agitation | 59 (26.3) | 40 (18.2) | 33 (16.3) | 19 (17.0) | 18 (18.8) | 13 (15.1) | 23 (32.9) | 6 (18.2) | 5 (21.7) | 3 (18.8) | 4 (26.7) | 2 (22.2) | 2 (20.0) | 1 (12.5) | 0 (0.0) |
| Appetite changes | 36 (16.1) | 49 (22.3) | 43 (21.3) | 15 (13.4) | 18 (18.8) | 28 (32.6) | 16 (22.9) | 7 (21.2) | 8 (34.8) | 2 (12.5) | 3 (20.0) | 1 (11.1) | 0 (0.0) | 0 (0.0) | 0 (0.0) |
| Attention/concentration difficulties | 42 (18.8) | 30 (13.6) | 38 (18.8) | 26 (23.2) | 21 (21.9) | 18 (20.9) | 8 (11.4) | 6 (18.2) | 2 (8.7) | 5 (31.2) | 5 (33.3) | 1 (11.1) | 1 (10.0) | 2 (25.0) | 0 (0.0) |
| Headaches | 24 (10.7) | 35 (15.9) | 25 (12.4) | 17 (15.2) | 17 (17.7) | 11 (12.8) | 11 (15.7) | 3 (9.1) | 4 (17.4) | 2 (12.5) | 3 (20.0) | 1 (11.1) | 0 (0.0) | 1 (12.5) | 0 (0.0) |
| Diarrhoea or constipation | 29 (12.9) | 33 (15.0) | 30 (14.9) | 13 (11.6) | 13 (13.5) | 6 (7.0) | 5 (7.1) | 6 (18.2) | 1 (4.3) | 2 (12.5) | 8 (53.3) | 2 (22.2) | 3 (30.0) | 1 (12.5) | 1 (50.0) |
| Feeling/being sick | 26 (11.6) | 37 (16.8) | 30 (14.9) | 10 (8.9) | 15 (15.6) | 4 (4.7) | 12 (17.1) | 5 (15.2) | 3 (13.0) | 2 (12.5) | 4 (26.7) | 1 (11.1) | 1 (10.0) | 1 (12.5) | 0 (0.0) |
| Memory problems | 20 (8.9) | 27 (12.3) | 22 (10.9) | 19 (17.0) | 15 (15.6) | 10 (11.6) | 6 (8.6) | 7 (21.2) | 1 (4.3) | 2 (12.5) | 7 (46.7) | 0 (0.0) | 1 (10.0) | 2 (25.0) | 0 (0.0) |
| Shaking | 26 (11.6) | 31 (14.1) | 26 (12.9) | 5 (4.5) | 14 (14.6) | 8 (9.3) | 4 (5.7) | 6 (18.2) | 3 (13.0) | 2 (12.5) | 2 (13.3) | 1 (11.1) | 1 (10.0) | 1 (12.5) | 0 (0.0) |
| Fast heartbeat | 16 (7.1) | 26 (11.8) | 22 (10.9) | 15 (13.4) | 8 (8.3) | 7 (8.1) | 7 (10.0) | 7 (21.2) | 0 (0.0) | 2 (12.5) | 4 (26.7) | 3 (33.3) | 3 (30.0) | 2 (25.0) | 0 (0.0) |
| Indigestion or stomach aches | 23 (10.3) | 27 (12.3) | 25 (12.4) | 9 (8.0) | 10 (10.4) | 5 (5.8) | 10 (14.3) | 4 (12.1) | 1 (4.3) | 1 (6.2) | 3 (20.0) | 0 (0.0) | 0 (0.0) | 1 (12.5) | 0 (0.0) |
| Muscle pain | 10 (4.5) | 12 (5.5) | 5 (2.5) | 7 (6.2) | 7 (7.3) | 5 (5.8) | 3 (4.3) | 4 (12.1) | 1 (4.3) | 1 (6.2) | 3 (20.0) | 0 (0.0) | 0 (0.0) | 0 (0.0) | 0 (0.0) |

| | | | | | | | | | | | | | | | |
|---|---|---|---|---|---|---|---|---|---|---|---|---|---|---|---|
| Itching/rash | 12 (5.4) | 10 (4.5) | 8 (4.0) | 10 (8.9) | 7 (7.3) | 1 (1.2) | 2 (2.9) | 0 (0.0) | 0 (0.0) | 0 (0.0) | 1 (6.7) | 0 (0.0) | 0 (0.0) | 2 (25.0) | 0 (0.0) |
| Blurred vision | 6 (2.7) | 7 (3.2) | 7 (3.5) | 7 (6.2) | 6 (6.2) | 3 (3.5) | 1 (1.4) | 5 (15.2) | 0 (0.0) | 1 (6.2) | 4 (26.7) | 2 (22.2) | 0 (0.0) | 1 (12.5) | 0 (0.0) |

**Table S11.** Prevalence of each side effect across antidepressant classes.

| Side Effect | Prevalence N (%) | | | |
|---|---|---|---|---|
| | SSRI | Atypical | SNRI | Tricyclic |
| Sleep disturbances | 223 (30.2) | 9 (50.0) | 45 (34.9) | 103 (42.9) |
| Weight gain/loss | 203 (27.5) | 4 (22.2) | 34 (26.4) | 93 (38.8) |
| Nausea | 223 (30.2) | 4 (22.2) | 42 (32.6) | 50 (20.8) |
| Fatigue | 183 (24.8) | 5 (27.8) | 32 (24.8) | 88 (36.7) |
| Dry mouth | 168 (22.7) | 15 (83.3) | 40 (31.0) | 71 (29.6) |
| Sexual function or interest | 236 (31.9) | 1 (5.6) | 30 (23.3) | 27 (11.2) |
| Dizziness | 155 (21.0) | 4 (22.2) | 37 (28.7) | 55 (22.9) |
| Increased anxiety/agitation | 160 (21.7) | 3 (16.7) | 24 (18.6) | 41 (17.1) |
| Appetite changes | 152 (20.6) | 0 (0.0) | 25 (19.4) | 49 (20.4) |
| Attention/concentration difficulties | 120 (16.2) | 3 (16.7) | 27 (20.9) | 55 (22.9) |
| Headaches | 99 (13.4) | 1 (5.6) | 20 (15.5) | 34 (14.2) |
| Diarrhoea or constipation | 98 (13.3) | 4 (22.2) | 19 (14.7) | 32 (13.3) |
| Feeling/being sick | 108 (14.6) | 2 (11.1) | 20 (15.5) | 21 (8.8) |
| Memory problems | 76 (10.3) | 3 (16.7) | 22 (17.1) | 38 (15.8) |
| Shaking | 90 (12.2) | 2 (11.1) | 20 (15.5) | 18 (7.5) |
| Fast heartbeat | 71 (9.6) | 5 (27.8) | 15 (11.6) | 31 (12.9) |
| Indigestion or stomach aches | 86 (11.6) | 1 (5.6) | 14 (10.9) | 18 (7.5) |
| Muscle pain | 31 (4.2) | 0 (0.0) | 11 (8.5) | 16 (6.7) |
| Itching/rash | 32 (4.3) | 2 (11.1) | 7 (5.4) | 12 (5.0) |
| Blurred vision | 21 (2.8) | 1 (5.6) | 11 (8.5) | 17 (7.1) |

**Table S12.** Number of users endorsing at least one side effect by antidepressant class.

| Class | Users N | Users endorsing side effects N (%) |
|---|---|---|
| SSRI | 1769 | 674 (38.1%) |
| Atypical | 228 | 97 (42.5%) |
| SNRI | 294 | 113 (38.4%) |
| Tricyclic | 383 | 144 (37.6%) |

**Table S13.** PHQ-9 and GAD-7 scores in responders and non-responders.

| | N (%) | |
|---|---|---|
| | Responders | Non-Responders |
| **PHQ-9 score, mean (SD)** | 6.24 (5.21) | 13.11 (9.12) |
| Minimal (0-4) | 57 (44.9) | 3 (33.3) |
| Mild (5-9) | 43 (33.9) | 0 (0) |
| Moderate (10-14) | 16 (12.6) | 1 (11.1) |
| Moderately Severe (15-19) | 8 (6.3) | 3 (33.3) |
| Severe (20-27) | 3 (2.4) | 2 (22.2) |
| **GAD-7 score, mean (SD)** | 4.73 (4.45) | 11.89 (5.46) |
| Minimal (0-4) | 71 (55.5) | 1 (11.1) |
| Mild (5-9) | 42 (32.8) | 1 (11.1) |
| Moderate (10-14) | 10 (7.8) | 3 (33.3) |
| Severe (15-21) | 5 (3.9) | 4 (44.4) |

**Table S14.** Top 20 most endorsed depression symptoms and their average impact in the responder group.

| Domain | Symptom | N (%) | Impact Mean (SD) |
|---|---|---|---|
| Physical | Lack of energy | 270 (96.4) | 3.48 (0.88) |
| Sleep | Waking unrefreshed/always fatigued | 266 (95.0) | 3.61 (0.91) |
| Behaviour | Reduced social activity | 255 (91.7) | 3.16 (0.93) |
| Depression | Loss of interest in hobbies | 245 (90.4) | 3.32 (0.89) |
| Anxiety | Anxious/worrying | 242 (86.4) | 3.37 (0.89) |
| Behaviour | Sensitive to criticism or rejection | 240 (86.0) | 3.40 (0.99) |
| Depression | Depressed mood: sadness, hopelessness, frequent crying | 237 (85.6) | 3.39 (0.84) |
| Cognitive Function | Everything feels like a burden | 237 (84.6) | 3.45 (0.91) |
| Cognitive Function | Racing thoughts | 234 (83.6) | 3.58 (0.95) |
| Sleep | Waking throughout night | 224 (80.0) | 3.33 (0.83) |
| Behaviour | Loss of interest in sex | 219 (79.3) | 2.91 (1.14) |
| Depression | Worthlessness and self-doubt | 216 (78.5) | 3.42 (0.89) |
| Cognitive Function | Can't concentrate on book/movie | 216 (77.4) | 3.00 (0.89) |
| Cognitive Function | Can't concentrate at work/study | 216 (77.1) | 3.43 (0.89) |
| Cognitive Function | Reduced ability to think | 215 (77.1) | 3.25 (0.87) |
| Cognitive Function | Decreased creativity | 210 (75.8) | 2.82 (0.95) |
| Sleep | Difficulty getting to sleep | 208 (74.3) | 3.52 (0.86) |
| Physical | Low sex drive | 207 (74.5) | 2.68 (1.05) |
| Depression | Guilt: blaming yourself | 204 (74.7) | 3.24 (0.85) |
| Behaviour | Reduced daytime activity | 202 (72.9) | 3.18 (0.85) |

**Table S15.** Top 20 most endorsed depression symptoms and their average impact in the non-responder group.

| Domain | Symptom | N (%) | Impact Mean (SD) |
|---|---|---|---|
| Depression | Depressed mood: sadness, hopelessness, frequent crying | 18 (100.0) | 3.44 (0.70) |
| Cognitive Function | Racing thoughts | 18 (100.0) | 3.67 (0.97) |
| Physical | Lack of energy | 18 (100.0) | 3.67 (0.77) |
| Depression | Worthlessness and self-doubt | 17 (94.4) | 3.88 (0.93) |
| Sleep | Waking unrefreshed | 17 (94.4) | 3.88 (0.78) |
| Behaviour | Sensitive to criticism or rejection | 17 (94.4) | 3.53 (0.94) |
| Depression | Guilt: blaming yourself | 16 (88.9) | 3.19 (0.83) |
| Cognitive Function | Everything feels like a burden | 16 (88.9) | 3.75 (0.86) |
| Cognitive Function | Reduced ability to think | 15 (83.3) | 3.67 (0.72) |
| Behaviour | Reduced social activity | 15 (83.3) | 3.87 (0.92) |
| Anxiety | Anxious/worrying | 14 (77.8) | 3.64 (0.74) |
| Anxiety | Avoidance of public areas | 14 (77.8) | 4.14 (1.03) |
| Anxiety | Persistent focus on past trauma | 14 (77.8) | 3.57 (1.02) |
| Cognitive Function | Can't concentrate on book/movie | 14 (77.8) | 3.00 (0.88) |
| Cognitive Function | Reduced ability to make decisions | 14 (77.8) | 3.43 (0.85) |
| Cognitive Function | Decreased creativity | 14 (77.8) | 2.93 (0.92) |
| Sleep | Difficulty getting to sleep | 14 (77.8) | 4.07 (0.73) |
| Behaviour | Reduced daytime activity | 14 (77.8) | 3.43 (0.85) |
| Depression | Loss of interest in hobbies | 13 (72.2) | 3.54 (0.97) |
| Sleep | Waking throughout night | 13 (72.2) | 3.92 (0.95) |

**Table S16.** Antidepressant usage among responders and non-responders.

| Antidepressant | Class | N (%) | |
|---|---|---|---|
| | | Responders | Non-responders |
| Citalopram | SSRI | 96 (34.2) | 8 (44.4) |
| Paroxetine | SSRI | 25 (8.9) | 5 (27.8) |
| Escitalopram | SSRI | 10 (3.6) | 1 (5.6) |
| Fluoxetine | SSRI | 90 (32.0) | 13 (72.2) |
| Sertraline | SSRI | 101 (35.9) | 15 (83.3) |
| Venlafaxine | SNRI | 14 (5.0) | 6 (33.3) |
| Duloxetine | SNRI | 5 (1.8) | 3 (16.7) |
| Amitriptyline | Tricyclic | 19 (6.8) | 2 (11.1) |
| Mirtazapine | Atypical | 21 (7.5) | 7 (38.9) |
| Trazodone | Atypical | 1 (0.4) | 0 (0.0) |
| Clomipramine | Tricyclic | 2 (0.7) | 1 (5.6) |
| Dosulepin | Tricyclic | 2 (0.7) | 0 (0.0) |
| Nortriptyline | Tricyclic | 1 (0.4) | 0 (0.0) |
| Lofepramine | Tricyclic | 0 (0.0) | 0 (0.0) |

| | | | |
|---|---|---|---|
| Imipramine | Tricyclic | 1 (0.4) | 2 (11.1) |

**Table S17.** Efficacy ratings among responders for each antidepressant.

| Antidepressant | Number of users N | N (%) | |
|---|---|---|---|
| | | Yes, a lot | Yes, a little |
| Citalopram | 93 | 82 (88.2) | 11 (11.8) |
| Paroxetine | 24 | 20 (83.3) | 4 (16.7) |
| Escitalopram | 10 | 7 (70.0) | 3 (30.0) |
| Fluoxetine | 89 | 81 (91.0) | 8 (9.0) |
| Sertraline | 101 | 94 (93.1) | 7 (6.9) |
| Venlafaxine | 13 | 13 (100.0) | 0 (0.0) |
| Duloxetine | 5 | 4 (80.0) | 1 (20.0) |
| Amitriptyline | 16 | 10 (62.5) | 6 (37.5) |
| Mirtazapine | 19 | 16 (84.2) | 3 (15.8) |
| Clomipramine | 1 | 1 (100.0) | 0 (0.0) |
| Imipramine | 1 | 0 (0.0) | 1 (100.0) |

Table S18. Prevalence of side effects in responder and non-responder groups.

| Side Effect | N (%) | |
|---|---|---|
| | Responders | Non-responders |
| Sleep disturbances | 37 (22.3) | 6 (26.1) |
| Weight gain/loss | 46 (27.7) | 5 (21.7) |
| Nausea | 49 (29.5) | 2 (8.7) |
| Fatigue/lack of energy | 29 (17.5) | 5 (21.7) |
| Dry mouth | 47 (28.3) | 2 (8.7) |
| Sexual function or interest | 67 (40.4) | 6 (26.1) |
| Dizziness | 29 (17.5) | 6 (26.1) |
| Increased anxiety/agitation | 14 (8.4) | 5 (21.7) |
| Appetite changes | 32 (19.3) | 3 (13.0) |
| Attention/concentration difficulties | 13 (7.8) | 3 (13.0) |
| Headaches | 17 (10.2) | 3 (13.0) |
| Diarrhoea or constipation | 24 (14.5) | 1 (4.3) |
| Feeling/being sick | 8 (4.8) | 1 (4.3) |
| Memory problems | 7 (4.2) | 1 (4.3) |
| Shaking | 14 (8.4) | 4 (17.4) |
| Fast heartbeat | 13 (7.8) | 2 (8.7) |
| Indigestion or stomach aches | 12 (7.2) | 3 (13.0) |
| Muscle pain | 3 (1.8) | 2 (8.7) |
| Itching/rash | 3 (1.8) | 2 (8.7) |
| Blurred vision | 6 (3.6) | 0 (0.0) |

**Table S19.** PHQ-9 and GAD-7 scores in SNRI responders.

| | N (%) |
|---|---|
| **PHQ-9 score, mean (SD)** | 16.38 (6.59) |
| Minimal (0-4) | 1 (12.5) |
| Mild (5-9) | 2 (25.0) |
| Moderate (10-14) | 2 (25.0) |
| Moderately Severe (15-19) | 3 (37.5) |
| Severe (20-27) | 1 (12.5) |
| **GAD-7 score, mean (SD)** | 13.88 (5.30) |
| Minimal (0-4) | 1 (12.5) |
| Mild (5-9) | 1 (12.5) |
| Moderate (10-14) | 1 (12.5) |
| Severe (15-21) | 5 (62.5) |

**Table S20.** Top 20 most endorsed depression symptoms and their average impact on daily life in the SNRI responder group. Impact rated on a 5-point scale: 1 (no impact) to 5 (severe impact).

| Domain | Symptom | N (%) | Impact Mean (SD) |
|---|---|---|---|
| Depression | Depressed mood: sadness, hopelessness, frequent crying | 9 (100.0) | 3.89 (0.93) |
| Depression | Guilt: blaming yourself | 9 (100.0) | 3.56 (1.24) |
| Depression | Worthlessness and self-doubt | 9 (100.0) | 4.00 (1.00) |
| Depression | Loss of interest in hobbies | 9 (100.0) | 3.56 (1.01) |
| Depression | Suicidal feelings or thoughts | 9 (100.0) | 3.56 (1.33) |
| Anxiety | Avoidance of public areas | 9 (100.0) | 4.56 (0.53) |
| Cognitive Function | Everything feels like a burden | 9 (100.0) | 3.89 (0.93) |
| Cognitive Function | Reduced ability to make decisions | 9 (100.0) | 3.56 (1.13) |
| Sleep | Waking unrefreshed | 9 (100.0) | 4.11 (0.78) |
| Behaviour | Reduced daytime activity | 9 (100.0) | 3.67 (0.71) |
| Behaviour | Loss of interest in sex | 9 (100.0) | 3.78 (1.48) |
| Physical | Lack of energy | 9 (100.0) | 4.44 (0.53) |
| Physical | Headaches/migraines | 9 (100.0) | 3.33 (0.87) |
| Depression | Feeling detached from reality | 8 (88.9) | 3.50 (1.20) |
| Depression | No feelings of affection | 8 (88.9) | 3.62 (0.74) |
| Cognitive Function | Can't concentrate on book/movie | 8 (88.9) | 3.12 (0.99) |
| Cognitive Function | Can't concentrate at work/study | 8 (88.9) | 4.00 (0.76) |
| Cognitive Function | Can't complete everyday tasks | 8 (88.9) | 4.50 (0.53) |
| Sleep | Waking throughout night | 8 (88.9) | 3.75 (0.89) |
| Behaviour | Sensitive to criticism or rejection | 8 (88.9) | 3.62 (1.06) |

**Table S21.** Antidepressant usage and efficacy ratings in the SNRI responder group.

| Antidepressant | Class | N (%) | | | |
|---|---|---|---|---|---|
| | | Usage | Yes, a lot | Yes, a little | No |
| Venlafaxine | SNRI | 9 | 4 (44.4) | 2 (22.2) | 3 (33.3) |
| Citalopram | SSRI | 7 | 0 (0.0) | 0 (0.0) | 7 (100.0) |
| Fluoxetine | SSRI | 7 | 0 (0.0) | 0 (0.0) | 7 (100.0) |
| Sertraline | SSRI | 7 | 0 (0.0) | 0 (0.0) | 7 (100.0) |
| Paroxetine | SSRI | 3 | 0 (0.0) | 0 (0.0) | 3 (100.0) |
| Duloxetine | SNRI | 3 | 0 (0.0) | 3 (100.0) | 0 (0.0) |
| Amitriptyline | Tricyclic | 3 | 0 (0.0) | 0 (0.0) | 3 (100.0) |
| Mirtazapine | Atypical | 3 | 1 (33.3) | 1 (33.3) | 1 (33.3) |
| Trazodone | Atypical | 3 | 0 (0.0) | 1 (33.3) | 2 (66.7) |
| Clomipramine | Tricyclic | 2 | 1 (50.0) | 1 (50.0) | 0 (0.0) |
| Nortriptyline | Tricyclic | 2 | 1 (50.0) | 0 (0.0) | 1 (50.0) |
| Lofepramine | Tricyclic | 1 | 1 (100.0) | 0 (0.0) | 0 (0.0) |

**Table S22.** Prevalence of side effects in the SNRI responder group.

| Side Effect | N (%) |
|---|---|
| Dizziness | 9 (64.3) |
| Sleep disturbances | 9 (64.3) |
| Nausea | 7 (50.0) |
| Feeling sick | 6 (42.9) |
| Increased anxiety | 6 (42.9) |
| Sexual function or interest | 6 (42.9) |
| Fatigue | 5 (35.7) |
| Shaking | 5 (35.7) |
| Weight gain/loss | 5 (35.7) |
| Dry mouth | 4 (28.6) |
| Fast heartbeat | 4 (28.6) |
| Bowel issues | 3 (21.4) |
| Appetite changes | 1 (7.1) |
| Concentration difficulties | 1 (7.1) |
| Memory problems | 1 (7.1) |
| Blurred vision | 0 (0.0) |
| Headaches | 0 (0.0) |
| Muscle pain | 0 (0.0) |
| Rash | 0 (0.0) |
| Stomach issues | 0 (0.0) |
