## Supplementary material for "Cohort Profile: Investigating Antidepressant Response within Generation Scotland": Questionnaire


Megan Calnan[1], Amelia Edmondson – Stait[1], Hannah Milbourn[2], Esme Elsden[1], Anjali K. Henders[4], Emily L. Ball[1], Matthew H. Iveson[1], AMBER Research Team, AMBER Lived Experience Advisory Panel, Generation Scotland Team, Naomi R. Wray[3,4], Sonia Shah[4], Cathryn M. Lewis[5,6], and Andrew M. McIntosh[1]

1. Institute for Neuroscience and Cardiovascular Research, University of Edinburgh, Edinburgh, United Kingdom
2. Centre for Genomic and Experimental Medicine, Institute of Genetics and Cancer, University of Edinburgh, Edinburgh, United Kingdom
3. Department of Psychiatry, University of Oxford, United Kingdom
4. Institute for Molecular Bioscience, University of Queensland, Brisbane, Queensland, Australia
5. Social, Genetic & Developmental Psychiatry Centre, King's College London, London, United Kingdom
6. Department of Medical and Molecular Genetics, King's College London, London, United Kingdom


This questionnaire was adapted from the instrument developed in the Cell-omics Resource of the Australian Genetics of Depression Study (AGDS:Cell-o) (Mitchell et al., 2025).


Mitchell, B., Gilroy, D., Wallace, L., Kiewa, J., Parker, R., Nunn, L., Walker, A., Lin, T., Ziser, L., English, G., O'Neill, M., Pertile, R., McKenna, S., Hockey, S., Hill, A., Shim, A., To, T., Treneman, A., McIntosh, A., … Wray, N. (2025). Cohort profile: Cell-omics Resource of the Australian Genetics of Depression Study (AGDS:Cell-o). https://doi.org/10.1101/2025.09.16.25335873

Thank for agreeing to complete the questionnaire.  If you are unable to complete the questionnaire by yourself, you can ask a carer or family member to assist you. If you feel you need any further help with this questionnaire, please reach out the Research Assistant, Megan Calnan at. Please let us know who will be completing the questionnaire.

◯ I am completing this myself

◯ I have help from a carer or family member

◯ The Research Assistant is helping me

**Have you ever taken any of the following medications to treat your mood?**

- Citalopram or Celexa
- Paroxetine or Seroxat
- Escitalopram or Lexapro
- Fluoxetine or Prozac
- Sertraline or Lustral
- Venlafaxine or Efexor
- Duloxetine or Cymbalta
- Amitriptyline or Elavil
- Mirtazapine or Remeron
- Trazodone or Molipaxin
- Clomipramine or Anafranil
- Dosulepin or Prothiaden
- Nortriptyline or Pamelor
- Lofepramine or Lomont
- Imipramine or Tofranil

◯ Yes

◯ No

**Section 1: Your Depression**

In this section, we want to understand how you feel when you are depressed and struggling with your depression (an episode of depression). Each person has a unique experience with depression and how it makes them feel. Symptoms associated with depression can be grouped into 6 categories: sleep and body clock, behavioural, physical, mood, anxiety, and cognitive functioning symptoms.

When you are feeling depressed, do you experience any of the following symptoms related to your **mood**? Please answer YES or NO to each of the symptoms listed.

| | YES | NO |
|---|---|---|
| Depression Mood; sadness, hopelessness, frequent crying | ○ | ○ |
| Feelings of Guilt: blaming yourself and feel you are failing | ○ | ○ |
| Feelings of worthlessness and self-doubt | ○ | ○ |
| Loss of interest in hobbies and in things you usually enjoyed | ○ | ○ |
| I am feeling detached from reality | ○ | ○ |
| Don’t have feelings of affection even when you know you should | ○ | ○ |
| Surroundings feel unreal or like looking through a thick fog | ○ | ○ |
| Suicidal feelings or thinking about suicide | ○ | ○ |

Rate the impact of these symptoms on your day-to-day life when you are in a depressive episode.

| | No Impact | Slight Impact | Moderate Impact | Major Impact | Severe Impact |
|---|---|---|---|---|---|
| Depression Mood; sadness, hopelessness, frequent crying | ○ | ○ | ○ | ○ | ○ |
| Feelings of Guilt: blaming yourself and feel you are failing | ○ | ○ | ○ | ○ | ○ |
| Feelings of worthlessness and self-doubt | ○ | ○ | ○ | ○ | ○ |
| Loss of interest in hobbies and in things you usually enjoyed | ○ | ○ | ○ | ○ | ○ |
| Feeling detached from reality | ○ | ○ | ○ | ○ | ○ |
| Don’t have feelings of affection even when you know you should | ○ | ○ | ○ | ○ | ○ |
| Surroundings feel unreal or in a thick fog | ○ | ○ | ○ | ○ | ○ |
| Suicidal feelings or thinking about suicide | ○ | ○ | ○ | ○ | ○ |

When you are feeling depressed, do you experience any of the following symptoms related to **anxiety**? Please answer YES or NO to each of the symptoms listed.

| | YES | NO |
|---|---|---|
| Anxious – worrying about minor things | ○ | ○ |
| Repetitive fearful thoughts - like being afraid of catching an infection, or that a disaster might happen | ○ | ○ |
| Feeling suspicious of other people | ○ | ○ |
| Intrusive, recurrent thoughts about your own health | ○ | ○ |
| Engagement in very repetitive behaviours that are linked to your anxiety - like unnecessary washing, cleaning, or checking things over | ○ | ○ |
| Panic attacks | ○ | ○ |
| Avoidance of public areas going out in public or travelling outside your home | ○ | ○ |
| Persistent focus on past traumatic events | ○ | ○ |

Rate the impact of these symptoms on your day-to-day life when you are in a depressive episode.

| | No Impact | Slight Impact | Moderate Impact | Major Impact | Severe Impact |
|---|---|---|---|---|---|
| Anxious – worrying about minor things | ○ | ○ | ○ | ○ | ○ |
| Repetitive fearful thoughts - like being afraid of catching an infection, or that a disaster might happen | ○ | ○ | ○ | ○ | ○ |
| Feeling suspicious of other people | ○ | ○ | ○ | ○ | ○ |
| Intrusive, recurrent thoughts about your own health | ○ | ○ | ○ | ○ | ○ |
| Engagement in very repetitive behaviours that are linked to your anxiety - like unnecessary washing, cleaning, or checking things over | ○ | ○ | ○ | ○ | ○ |
| Panic attacks | ○ | ○ | ○ | ○ | ○ |
| Avoidance of public areas going out in public or travelling outside your home | ○ | ○ | ○ | ○ | ○ |
| Persistent focus on past traumatic events | ○ | ○ | ○ | ○ | ○ |

When you are feeling depressed, do you experience any of the following symptoms related to your **memory, ability to focus and think (general cognitive function)**? Please answer YES or NO to each of the symptoms listed.

| | YES | NO |
|---|---|---|
| Reduced ability to think and process thoughts | ○ | ○ |
| Everything feels like it is a burden: can't be bothered with anything | ○ | ○ |
| Reduced ability to concentrate on a book or movie | ○ | ○ |
| Reduced ability to concentrate at work/study | ○ | ○ |
| Reduced ability to complete everyday tasks | ○ | ○ |
| Reduced ability to manage the workload expected of you | ○ | ○ |
| Racing thoughts - Your mind isn't able to "shut off" and you can't fully relax | ○ | ○ |
| Reduced ability to make decisions | ○ | ○ |
| Decreased creativity | ○ | ○ |
| Increased ability to focus and manage the workload expected of you | ○ | ○ |

Rate the impact of these symptoms on your day-to-day life when you are in a depressive episode.

| | No Impact | Slight Impact | Moderate Impact | Major Impact | Severe Impact |
|---|---|---|---|---|---|
| Reduced ability to think and process thoughts | ○ | ○ | ○ | ○ | ○ |
| Everything feels like it is a burden: can't be bothered with anything | ○ | ○ | ○ | ○ | ○ |
| Reduced ability to concentrate on a book or movie | ○ | ○ | ○ | ○ | ○ |
| Reduced ability to concentrate at work/study | ○ | ○ | ○ | ○ | ○ |
| Reduced ability to complete everyday tasks | ○ | ○ | ○ | ○ | ○ |
| Reduced ability to manage the workload expected of you | ○ | ○ | ○ | ○ | ○ |
| Racing thoughts - Your mind isn't able to "shut off" and you can't fully relax | ○ | ○ | ○ | ○ | ○ |
| Reduced ability to make decisions | ○ | ○ | ○ | ○ | ○ |
| Decreased creativity | ○ | ○ | ○ | ○ | ○ |
| Increased ability to focus and manage the workload expected of you | ○ | ○ | ○ | ○ | ○ |

When you are feeling depressed, do you experience any of the following changes in your **sleep pattern**? Please answer YES or NO to each of the symptoms listed.

| | YES | NO |
|---|---|---|
| Difficulty getting to sleep | ○ | ○ |
| Waking throughout the night | ○ | ○ |
| Waking up much later | ○ | ○ |
| Oversleeping (sleep more than 8-10 hours a night) | ○ | ○ |
| Waking up feeling unrefreshed, always feeling fatigued | ○ | ○ |
| Feel worst in the morning | ○ | ○ |
| Internal body clock seems out of sync with actual time of day - like jetlag | ○ | ○ |
| Early morning (before sunrise) wakening | ○ | ○ |

Rate the impact of these symptoms on your day-to-day life when you are in a depressive episode.

| | No Impact | Slight Impact | Moderate Impact | Major Impact | Severe Impact |
|---|---|---|---|---|---|
| Difficulty getting to sleep | ○ | ○ | ○ | ○ | ○ |
| Waking throughout the night | ○ | ○ | ○ | ○ | ○ |
| Waking up much later | ○ | ○ | ○ | ○ | ○ |
| Oversleeping (sleep more than 8-10 hours a night) | ○ | ○ | ○ | ○ | ○ |
| Waking up feeling unrefreshed, always feeling fatigued | ○ | ○ | ○ | ○ | ○ |
| Feel worst in the morning | ○ | ○ | ○ | ○ | ○ |
| Internal body clock seems out of sync with actual time of day - like jetlag | ○ | ○ | ○ | ○ | ○ |
| Early morning (before sunrise) wakening | ○ | ○ | ○ | ○ | ○ |

When you are feeling depressed, do you experience any of the following changes in your **behaviour**? Please answer YES or NO to each of the symptoms listed.

| | YES | NO |
|---|---|---|
| Unable to sit still, feeling the need to keep moving | ○ | ○ |
| Reduced daytime activity | ○ | ○ |
| Increased sensitivity to criticism or rejection | ○ | ○ |
| Spend money without thinking it through | ○ | ○ |
| Easily irritated or frustrated - even unprovoked rage | ○ | ○ |
| Alienate friends and/or family | ○ | ○ |
| Reduced interest in participating in social activities | ○ | ○ |
| Loss of interest in sex | ○ | ○ |
| Engage very repetitive behaviours - like unnecessary washing, cleaning or checking over and over | ○ | ○ |
| Reluctance to talk: give single word answers when asked a question | ○ | ○ |

Rate the impact of these symptoms on your day-to-day life when you are in a depressive episode.

| | No Impact | Slight Impact | Moderate Impact | Major Impact | Severe Impact |
|---|---|---|---|---|---|
| Unable to sit still, feeling the need to keep moving | ○ | ○ | ○ | ○ | ○ |
| Reduced daytime activity | ○ | ○ | ○ | ○ | ○ |
| Increased sensitivity to criticism or rejection | ○ | ○ | ○ | ○ | ○ |
| Spend money without thinking it through | ○ | ○ | ○ | ○ | ○ |
| Easily irritated or frustrated - even unprovoked rage | ○ | ○ | ○ | ○ | ○ |
| Alienate friends and/or family | ○ | ○ | ○ | ○ | ○ |
| Reduced interest in participating in social activities | ○ | ○ | ○ | ○ | ○ |
| Loss of interest in sex | ○ | ○ | ○ | ○ | ○ |
| Engage very repetitive behaviours - like unnecessary washing, cleaning or checking over and over | ○ | ○ | ○ | ○ | ○ |
| Reluctance to talk: give single word answers when asked a question | ○ | ○ | ○ | ○ | ○ |

When you are feeling depressed, do you experience any of the following **physical symptoms**? Please answer YES or NO to each of the symptoms listed.

| | YES | NO |
|---|---|---|
| Lack of energy | ○ | ○ |
| Everything is in slow motion | ○ | ○ |
| Racing heart, sweat or have trouble breathing | ○ | ○ |
| Appetite increases | ○ | ○ |
| Appetite decreases | ○ | ○ |
| Significant weight gain | ○ | ○ |
| Significant weight loss | ○ | ○ |
| Heavy feeling in arms or legs | ○ | ○ |
| Unexplained aches and pains | ○ | ○ |
| Headaches and/or migraines | ○ | ○ |
| Gut problems e.g., constipation, peptic ulcers, reflux, irritable bowel | ○ | ○ |
| Low sex drive | ○ | ○ |
| Sexual dysfunction e.g., inability to get an erection or to reach orgasm or painful sex | ○ | ○ |
| Changes in menstrual cycle (female only, if applicable) | ○ | ○ |

Rate the impact of these symptoms on your day-to-day life when you are in a depressive episode.

| | No Impact | Slight Impact | Moderate Impact | Major Impact | Severe Impact |
|---|---|---|---|---|---|
| Lack of energy | ○ | ○ | ○ | ○ | ○ |
| Everything is in slow motion | ○ | ○ | ○ | ○ | ○ |
| Racing heart, sweat or have trouble breathing | ○ | ○ | ○ | ○ | ○ |
| Appetite increases | ○ | ○ | ○ | ○ | ○ |
| Appetite decreases | ○ | ○ | ○ | ○ | ○ |
| Significant weight gain | ○ | ○ | ○ | ○ | ○ |
| Significant weight loss | ○ | ○ | ○ | ○ | ○ |
| Heavy feeling in arms or legs | ○ | ○ | ○ | ○ | ○ |
| Unexplained aches and pains | ○ | ○ | ○ | ○ | ○ |
| Headaches and/or migraines | ○ | ○ | ○ | ○ | ○ |
| Gut problems e.g., constipation, peptic ulcers, reflux, irritable bowel | ○ | ○ | ○ | ○ | ○ |
| Low sex drive | ○ | ○ | ○ | ○ | ○ |
| Sexual dysfunction e.g., inability to get an erection or to reach orgasm or painful sex | ○ | ○ | ○ | ○ | ○ |

| | | | | | |
|---|---|---|---|---|---|
| Changes in menstrual cycle (female only, if applicable) | ○ | ○ | ○ | ○ | ○ |

Do you experience challenges in maintaining your health and physical wellbeing due to symptoms of depression? Please choose any relevant options where depression impacts on your ability to manage that aspect of your wellbeing. Check all that apply

- ☐ Dental or oral hygiene (e.g., brushing, flossing, regular dental appointments)
- ☐ Food quality, adequate nutrition, and diet management
- ☐ Bodily hygiene (e.g., washing, hair care, regular showering or bathing, physical activity)
- ☐ Keeping a clean home or living space (e.g., daily chores)

Do you find that the time of year (season) impacts your depressive symptoms?

- ○ Yes
- ○ No
- ○ Don't know
- ○ Prefer not to answer

During which times of the year do you notice an increase in depression or low energy?

- ☐ Spring
- ☐ Summer
- ☐ Autumn
- ☐ Winter

**Section 2: Psychological or Physical Treatments for Depression** We understand that the effectiveness of treatments for depression varies for each individual and can include more than just medication. We are interested, from your experience, what has been the most effective psychological therapy and physical treatment, or combination of treatments, in managing your depression when you are feeling depressed. In a later section we will ask you about medications you have tried. Here, we focus on the psychological therapy and/or physical treatment that has been most effective in relieving your symptoms of depression. For some people, and for a variety of reasons, the best treatment for relieving symptoms may mean this treatment is not their current treatment of choice. Still, we ask you to focus on the treatment you think has been most effective with respect to symptoms of depression.

In your experience, **has any treatment you've tried for depression** been effective in reducing your symptoms of depression? If anything has helped, even a little, please choose yes. You will have the chance to give more detail on how much each treatment has positively impacted your depression.

- ◯ Yes
- ◯ No
- ◯ Don't know
- ◯ Prefer not to answer

In your experience, which **psychological therapy** and **physical treatment** OR **combination of these** that you have tried were at all effective in **reducing your symptoms of depression**? Check all that apply,  even if they provided only slight relief.

- [ ] Cognitive behavioural therapy (CBT)
- [ ] Acceptance and commitment therapy (ACT)
- [ ] Interpersonal therapy (IPT)
- [ ] Dialectical behaviour therapy (DBT)
- [ ] Behavioural therapy
- [ ] Counselling for specific issues/challenges
- [ ] Physical exercise (e.g. gym, yoga)
- [ ] Meditation
- [ ] Repetitive Transcranial Magnetic Stimulation (rTMS/TMS)
- [ ] Electroconvulsive therapy (ECT)
- [ ] Light therapy
- [ ] Online or self-management interventions (e.g. mindfulness apps like headspace)
- [ ] Other therapy: ____________________________________________

Please rate the positive impact of each psychological therapies and physical treatments in reducing your symptoms of depression.

| | None | Minor Impact | Some Impact | Moderate Impact | Major Impact | Very Significant Impact |
|---|---|---|---|---|---|---|
| Cognitive behavioural therapy (CBT) | ○ | ○ | ○ | ○ | ○ | ○ |
| Acceptance and commitment therapy (ACT) | ○ | ○ | ○ | ○ | ○ | ○ |
| Interpersonal therapy (IPT) | ○ | ○ | ○ | ○ | ○ | ○ |
| Dialectical behaviour therapy (DBT) | ○ | ○ | ○ | ○ | ○ | ○ |
| Behavioural therapy | ○ | ○ | ○ | ○ | ○ | ○ |
| Counselling for specific issues/challenges | ○ | ○ | ○ | ○ | ○ | ○ |
| Physical exercise (e.g. gym, yoga) | ○ | ○ | ○ | ○ | ○ | ○ |
| Meditation | ○ | ○ | ○ | ○ | ○ | ○ |
| Repetitive Transcranial Magnetic Stimulation (rTMS/TMS) | ○ | ○ | ○ | ○ | ○ | ○ |
| Electroconvulsive therapy (ECT) | ○ | ○ | ○ | ○ | ○ | ○ |
| Light therapy | ○ | ○ | ○ | ○ | ○ | ○ |
| Online or self-management interventions (e.g. mindfulness apps like headspace) | ○ | ○ | ○ | ○ | ○ | ○ |
| Other therapy: | ○ | ○ | ○ | ○ | ○ | ○ |

Were any of the psychological therapies or physical treatments you selected received while you were taking antidepressant medication?

- ◯ Yes
- ◯ No
- ◯ Don't know
- ◯ Prefer not to answer

**Section 3: Medication Treatment for Depression** In this section of the questionnaire, we will be asking questions about any antidepressant medications you have take or are currently taking to manage your depression.
What medications have you **ever been prescribed but are not currently taking**?

- ☐ Citalopram or Celexa
- ☐ Paroxetine or Seroxat
- ☐ Escitalopram or Lexapro
- ☐ Fluoxetine or Prozac
- ☐ Sertraline or Lustral
- ☐ Venlafaxine or Efexor
- ☐ Duloxetine or Cymbalta
- ☐ Amitriptyline or Elavil
- ☐ Mirtazapine or Remeron
- ☐ Trazodone or Molipaxin
- ☐ Clomipramine or Anafranil
- ☐ Dosulepin or Prothiaden
- ☐ Nortriptyline or Pamelor
- ☐ Lofepramine or Lomont
- ☐ Imipramine or Tofranil

☐ Other: (please do not enter any personal identifying information)

____________________________________________

☐ I am currently prescribed antidepressant medication and have not stopped taking it

What prescribed medications have you **taken in the last year** to manage your depression?

- [ ] Citalopram or Celexa
- [ ] Paroxetine or Seroxat
- [ ] Escitalopram or Lexapro
- [ ] Fluoxetine or Prozac
- [ ] Sertraline or Lustral
- [ ] Venlafaxine or Efexor
- [ ] Duloxetine or Cymbalta
- [ ] Amitriptyline or Elavil
- [ ] Mirtazapine or Remeron
- [ ] Trazodone or Molipaxin
- [ ] Clomipramine or Anafranil
- [ ] Dosulepin or Prothiaden
- [ ] Nortriptyline or Pamelor
- [ ] Lofepramine or Lomont
- [ ] Imipramine or Tofranil
- [ ] Other: (please do not enter any personal identifying information)

________________________________________

☐ None

Why did you stop taking this medication? **We are displaying a list of the medication(s) you have tried but are not currently taking.**

| | I started feeling better | I experienced side effects | My symptoms were unchanged – it was not working for me | My prescription was changed | Other (please specify in next question): | Don’t know | Prefer not to answer |
|---|---|---|---|---|---|---|---|
| Medication(s) selected | ○ | ○ | ○ | ○ | ○ | ○ | ○ |

Q109 Please specify 'other' reason for stopping “medication name”: (please do not enter any personal identifying information)

________________________________________________________________

What prescribed medications are you **currently taking** to manage your depression?

- [ ] Citalopram or Celexa
- [ ] Paroxetine or Seroxat
- [ ] Escitalopram or Lexapro
- [ ] Fluoxetine or Prozac
- [ ] Sertraline or Lustral
- [ ] Venlafaxine or Efexor
- [ ] Duloxetine or Cymbalta
- [ ] Amitriptyline or Elavil
- [ ] Mirtazapine or Remeron
- [ ] Trazodone or Molipaxin
- [ ] Clomipramine or Anafranil
- [ ] Dosulepin or Prothiaden
- [ ] Nortriptyline or Pamelor
- [ ] Lofepramine or Lomont
- [ ] Imipramine or Tofranil
- [ ] Other: (please do not enter any personal identifying information)

____________________________________________

☐ None

How long did you take or have been taking this medication?

| | Days | Weeks | Months | Years |
|---|---|---|---|---|
| Medication(s) selected | | | | |

Please rate the impact of the medication(s) in reducing your symptoms of depression? (We are displaying a list of medications you are **NOT currently taking**)

| | None | Minor Impact | Some Impact | Moderate Impact | Major Impact | Extreme Impact |
|---|---|---|---|---|---|---|
| Medication(s) selected | ○ | ○ | ○ | ○ | ○ | ○ |

Please rate the impact of the medication(s) in reducing your symptoms of depression?(We are displaying a list of medications you **ARE currently taking**)

| | None | Minor Impact | Some Impact | Moderate Impact | Major Impact | Extreme Impact |
|---|---|---|---|---|---|---|
| Medication(s) selected | ○ | ○ | ○ | ○ | ○ | ○ |

Please rate the impact of the medication(s) that helped in improving the following areas of symptoms that you have reported you experience when feeling depressed?

| | None | Minor Impact | Some Impact | Moderate Impact | Major Impact | Significant Impact | Extreme Impact |
|---|---|---|---|---|---|---|---|
| Medication(s) selected: Sleep | ○ | ○ | ○ | ○ | ○ | ○ | ○ |
| Medication(s) selected: Behaviour | ○ | ○ | ○ | ○ | ○ | ○ | ○ |
| Medication(s) selected: Physical | ○ | ○ | ○ | ○ | ○ | ○ | ○ |
| Medication(s) selected: Mood | ○ | ○ | ○ | ○ | ○ | ○ | ○ |
| Medication(s) selected: Anxiety | ○ | ○ | ○ | ○ | ○ | ○ | ○ |
| Medication(s) selected: Cognitive functioning | ○ | ○ | ○ | ○ | ○ | ○ | ○ |

What was the **first area of symptoms** you noticed where the medication provided fast relief? (We are displaying a list of medications you are **NOT currently taking**)

| | None | Mood | Anxiety | Cognitive functioning | Sleep | Behaviour | Physical |
|---|---|---|---|---|---|---|---|
| Medication(s) selected | ○ | ○ | ○ | ○ | ○ | ○ | ○ |

What was the **first area of symptoms** you noticed where the medication provided fast relief? (We are displaying a list of medications you **ARE currently taking**)

| | None | Mood | Anxiety | Cognitive functioning | Sleep | Behaviour | Physical |
|---|---|---|---|---|---|---|---|
| Medication(s) selected | ○ | ○ | ○ | ○ | ○ | ○ | ○ |

How long did it take for you to notice any improvement in your symptoms after starting the medication(s)? (We are displaying a list of medications you are **NOT currently taking**)

| | 1-2 weeks | 2-4 weeks | 1-2 months | 2-4 months | 4-6 months | 6+ months | I didn't noticed any improvement | Don't know | Prefer not to answer |
|---|---|---|---|---|---|---|---|---|---|
| Medication(s) selected | ○ | ○ | ○ | ○ | ○ | ○ | ○ | ○ | ○ |

How long did it take for you to notice any improvement in your symptoms after starting the medication(s)? (We are displaying a list of medications you **ARE currently taking**)

| | 1-2 weeks | 2-4 weeks | 1-2 months | 2-4 months | 4-6 months | 6+ months | I haven't noticed any improvement | Don't know | Prefer not to answer |
|---|---|---|---|---|---|---|---|---|---|
| Medication(s) selected | ○ | ○ | ○ | ○ | ○ | ○ | ○ | ○ | ○ |

Did any of the medications below give fast relief from depression (within the first 2 weeks)?

- ☐ Citalopram or Celexa
- ☐ Paroxetine or Seroxat
- ☐ Escitalopram or Lexapro
- ☐ Fluoxetine or Prozac
- ☐ Sertraline or Lustral
- ☐ Venlafaxine or Efexor
- ☐ Duloxetine or Cymbalta
- ☐ Amitriptyline or Elavil
- ☐ Mirtazapine or Remeron
- ☐ Trazodone or Molipaxin
- ☐ Clomipramine or Anafranil
- ☐ Dosulepin or Prothiaden
- ☐ Nortriptyline or Pamelor
- ☐ Lofepramine or Lomont
- ☐ Imipramine or Tofranil
- ☐ Other: (please do not enter any personal identifying information)

________________________________________

☐ None

In your experience, have you found a single medication **or** combination of medications successful for reducing your symptoms of depression?

- ◯ Single medication
- ◯ Combination of medications
- ◯ None
- ◯ Don't know
- ◯ Prefer not to answer

In your experience, what has been the most effective **single medication** for reducing your symptoms of depression?

- ◯ Citalopram or Celexa
- ◯ Paroxetine or Seroxat
- ◯ Escitalopram or Lexapro
- ◯ Fluoxetine or Prozac
- ◯ Sertraline or Lustral
- ◯ Venlafaxine or Efexor
- ◯ Duloxetine or Cymbalta
- ◯ Amitriptyline or Elavil
- ◯ Mirtazapine or Remeron
- ◯ Trazodone or Molipaxin
- ◯ Clomipramine or Anafranil
- ◯ Dosulepin or Prothiaden
- ◯ Nortriptyline or Pamelor
- ◯ Lofepramine or Lomont

○ Imipramine or Tofranil

○ Other: (please do not enter any personal identifying information)

______________________________________________________

In your experience, what has been the most effective **combinations of medications** for reducing your symptoms of depression?

- ☐ Citalopram or Celexa
- ☐ Paroxetine or Seroxat
- ☐ Escitalopram or Lexapro
- ☐ Fluoxetine or Prozac
- ☐ Sertraline or Lustral
- ☐ Venlafaxine or Efexor
- ☐ Duloxetine or Cymbalta
- ☐ Amitriptyline or Elavil
- ☐ Mirtazapine or Remeron
- ☐ Trazodone or Molipaxin
- ☐ Clomipramine or Anafranil
- ☐ Dosulepin or Prothiaden
- ☐ Nortriptyline or Pamelor
- ☐ Lofepramine or Lomont
- ☐ Imipramine or Tofranil
- ☐ Other: (please do not enter any personal identifying information)

If you benefitted from a single medication (or combination of medications) for reducing your symptoms of depression, how quickly did it help you?

- ◯ Can't really specify
- ◯ 1-2 weeks
- ◯ 2-4 weeks
- ◯ 1-2 months
- ◯ 2-4 months
- ◯ 6+ months
- ◯ Don’t know
- ◯ Prefer not to answer

Overall, do you feel MEDICATION(S) SELECTED is effective in reducing your symptoms of depression?

- ◯ Yes, a lot
- ◯ Yes, a little
- ◯ No
- ◯ Don’t know
- ◯ Prefer not to answer

Q45 Have you used any of the following substances to try and manage your depression?

| | YES | NO |
|---|---|---|
| Alcohol | ○ | ○ |
| Tobacco | ○ | ○ |
| Cannabis | ○ | ○ |
| Stimulants | ○ | ○ |
| Other: (please do not enter any personal identifying information) | ○ | ○ |

**Section 4: Side Effects** In this section we will focus on side effects from any of the medications you have taken for depression.
Have you experienced any side effects from the medication(s)?

- Citalopram or Celexa
- Paroxetine or Seroxat
- Escitalopram or Lexapro
- Fluoxetine or Prozac
- Sertraline or Lustral
- Venlafaxine or Efexor
- Duloxetine or Cymbalta
- Amitriptyline or Elavil
- Mirtazapine or Remeron
- Trazodone or Molipaxin
- Clomipramine or Anafranil
- Dosulepin or Prothiaden
- Nortriptyline or Pamelor
- Lofepramine or Lomont
- Imipramine or Tofranil
- Other: (please do not enter any personal identifying information)

Can you please describe the side-effects you experienced with MEDICATION(S) SELECTED?

- ☐ Nausea
- ☐ Dizziness
- ☐ Sleep disturbances (insomnia or excessive sleepiness)
- ☐ Weight gain or loss
- ☐ Changes in appetite
- ☐ Fatigue, tired or lack of energy

- [ ] Increased anxiety or agitation
- [ ] Sexual function or interest
- [ ] Headaches
- [ ] Feeling and being sick
- [ ] Indigestion or stomach aches
- [ ] Diarrhoea or constipation
- [ ] Dry mouth
- [ ] Memory problems
- [ ] Attention/concentration difficulties
- [ ] Shaking
- [ ] Muscle pain
- [ ] Fast heartbeat
- [ ] Itching/rash
- [ ] Blurred vision
- [ ] Other: (please do not enter any personal identifying information)

________________________________________

On starting any of the medications you have ever taken, did you experience any side effects **within the first two weeks**?

- Citalopram or Celexa
- Paroxetine or Seroxat
- Escitalopram or Lexapro
- Fluoxetine or Prozac
- Sertraline or Lustral
- Venlafaxine or Efexor
- Duloxetine or Cymbalta
- Amitriptyline or Elavil
- Mirtazapine or Remeron
- Trazodone or Molipaxin
- Clomipramine or Anafranil
- Dosulepin or Prothiaden
- Nortriptyline or Pamelor
- Lofepramine or Lomont
- Imipramine or Tofranil
- Other: (please do not enter any personal identifying information)

you please describe the side-effects you experienced MEDICATION(S) SELECTED within the first two weeks?

- ☐ Nausea
- ☐ Dizziness
- ☐ Sleep disturbances (insomnia or excessive sleepiness)
- ☐ Weight gain or loss
- ☐ Changes in appetite
- ☐ Fatigue, tired or lack of energy
- ☐ Increased anxiety or agitation
- ☐ Sexual function or interest
- ☐ Headaches
- ☐ Feeling and being sick
- ☐ Indigestion or stomach aches
- ☐ Diarrhoea or constipation
- ☐ Dry mouth
- ☐ Memory problems
- ☐ Attention/concentration difficulties
- ☐ Shaking

- [ ] Muscle pain
- [ ] Fast heartbeat
- [ ] Itching/rash
- [ ] Blurred vision
- [ ] Other: (please do not enter any personal identifying information)
- [ ] Don’t know
- [ ] Prefer not to answer

Q84 What medications did you have to stop due to **severe side effects within the first two weeks**? Select all that apply.

- ☐ None
- ☐ Citalopram or Celexa
- ☐ Paroxetine or Seroxat
- ☐ Escitalopram or Lexapro
- ☐ Fluoxetine or Prozac
- ☐ Sertraline or Lustral
- ☐ Venlafaxine or Efexor
- ☐ Duloxetine or Cymbalta
- ☐ Amitriptyline or Elavil
- ☐ Mirtazapine or Remeron
- ☐ Trazodone or Molipaxin
- ☐ Clomipramine or Anafranil
- ☐ Dosulepin or Prothiaden
- ☐ Nortriptyline or Pamelor
- ☐ Lofepramine or Lomont
- ☐ Imipramine or Tofranil

- [ ] Other: (please do not enter any personal identifying information)

__________________________________________________________________

For any of the medications you have ever tried, did you experience any side effects **after the first month** of taking it?

- Citalopram or Celexa
- Paroxetine or Seroxat
- Escitalopram or Lexapro
- Fluoxetine or Prozac
- Sertraline or Lustral
- Venlafaxine or Efexor
- Duloxetine or Cymbalta
- Amitriptyline or Elavil
- Mirtazapine or Remeron
- Trazodone or Molipaxin
- Clomipramine or Anafranil
- Dosulepin or Prothiaden
- Nortriptyline or Pamelor
- Lofepramine or Lomont
- Imipramine or Tofranil
- Other: (please do not enter any personal identifying information)

Can you please describe the side-effects you experienced with MEDICATION(S) SELECTED after the first month?

- ☐ Nausea
- ☐ Dizziness
- ☐ Sleep disturbances (insomnia or excessive sleepiness)
- ☐ Weight gain or loss
- ☐ Changes in appetite
- ☐ Fatigue, tired or lack of energy
- ☐ Increased anxiety or agitation
- ☐ Sexual function or interest
- ☐ Headaches
- ☐ Feeling and being sick
- ☐ Indigestion or stomach aches
- ☐ Diarrhoea or constipation
- ☐ Dry mouth
- ☐ Memory problems
- ☐ Attention/concentration difficulties
- ☐ Shaking

- [ ] Muscle pain
- [ ] Fast heartbeat
- [ ] Itching/rash
- [ ] Blurred vision
- [ ] Other: (please do not enter any personal identifying information)
